# Model-based and model-free characterization of epidemic outbreaks

**DOI:** 10.1101/2020.09.16.20187484

**Authors:** Jonas Dehning, F. Paul Spitzner, Matthias C. Linden, Sebastian B. Mohr, Joao Pinheiro Neto, Johannes Zierenberg, Michael Wibral, Michael Wilczek, Viola Priesemann

**Author notes:** All authors contributed equally. Correspondence should be addressed to*.

## Abstract

Here, we provide detailed background information for our work on Bayesian inference of change-points in the spread of SARS-CoV-2 and the effectiveness of non-pharmaceutical interventions (Dehning et al., Science, 2020). We outline the general background of Bayesian inference and of SIR-like models. We explain the assumptions that underlie model-based estimates of the reproduction number and compare them to the assumptions that underlie model-free estimates, such as used in the Robert-Koch Institute situation reports. We highlight effects that originate from the two estimation approaches, and how they may cause differences in the inferred reproduction number. Furthermore, we explore the challenges that originate from data availability — such as publication delays and inconsistent testing — and explain their impact on the time-course of inferred case numbers. Along with alternative data sources, this allowed us to cross-check and verify our previous results.

## I. INTRODUCTION

After the release of our study “Inferring change points in the spread of COVID-19 reveals the effectiveness of interventions” in Science [1], we have received many constructive comments and interesting questions. Here, we address the most important of these questions and put them into perspective with respect to model-based and model free analyses of epidemic outbreaks. We provide additional background on our results, and we also discuss the robustness and performance of our published model in the light of newly available data.

The questions and comments we received can be broadly categorized into four topics:

1. Remarks on apparent discrepancies between the values for the estimated reproductive number 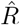 as reported by the Robert Koch Institute (RKI) and the corresponding spreading rate resulting from our published analysis. We will explain below how this apparent discrepancy partly arises from the comparison of model-free estimates to those from a differential-equation based modeling of disease dynamics. We show how the model-free approach may substantially underestimate the reproductive number *R* immediately after a sudden drop in *R* has occurred. From the comments we received it seems that this very important fact related to estimating *R*, i.e. 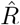, is largely unknown, and also counter-intuitive to most readers. This effect,together with the usage of alternative data (see point 3.), explains the apparent discrepancies between the RKI reports and our study. We therefore derive and demonstrate it in detail here.
2. Questions revolving around the philosophy and interpretation of our modeling approach that combines a differential equation model of the disease outbreak, Bayesian parameter inference and Bayesian model comparison. Most frequently we were asked if and in what sense our results have a causal interpretation. As we will explain below, our approach selects the most plausible of multiple causal explanations of the observed data, but does not establish strict interventional causality.
3. New data have been released in the time since our analyses were completed. Most prominently, data on the times of symptom onsets (epi curve) are now accessible in online data bases. The advantage is that the date of symptom onsets is closer to the infection date, allowing in principle a more precise estimation of the dynamics of the propagation. It brings however its owns source of errors, because the onset of symptoms is not reported for all cases, and the subjective reporting of symptom onset also bears some uncertainty. As we will show below, our central conclusions remain unchanged when updating our model to the new data.
4. Questions on how changes in testing capacity may have influenced our results. Given the data that have become available on the weekly (daily) number of performed tests, test capacity, and on delays between symptom onset, test and case report, we reanalyze in great detail the disease and testing dynamics, especially with respect to the timing of the peak in new symptom onsets. We conclude that all symptom onsets that are relevant for the main conclusions of our previous publication have been tested at a time when testing had sufficient capacity and was sufficiently constant.

We will in the following address the issues revolving around the reproductive number *R* first, also introducing the basic terminology of disease spreading and the fundamental difference between model-free and model-based estimation of epidemiological parameters. Next, we will discuss philosophy and interpretation of model-based estimation in the Bayesian framework and the causality question. We then show how our original analyses can be evolved to incorporate new data, in particular on symptom onset (epi curve). Last we turn to the important question of testing.

## II. ESTIMATING THE REPRODUCTIVE NUMBER *R*

### A. Basic SIR dynamics

Before we define the reproductive number *R*, we briefly recapitulate the basic SIR dynamics that we consider(Fig. 1). In principle, the course of an infection can be described as follows: A susceptible person (not infected and not immune) becomes infected but is initially not infectious; after some time, the person starts to be infectious but symptoms only show after the incubation period; eventually, the person is no longer infectious because she or he has been either isolated, has recovered, or died. The idea of compartmental models is to group the population into compartments; in the most simple but established SIR model these are susceptible (S), infected(I), and recovered (R). Assuming a well-mixed population (a mean-field approximation of everybody interacting with everybody), one can formulate differential equations that describe the time development of these compartments:

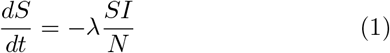

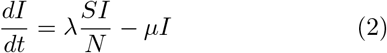

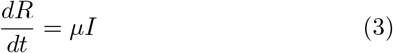

**FIG. 1.**
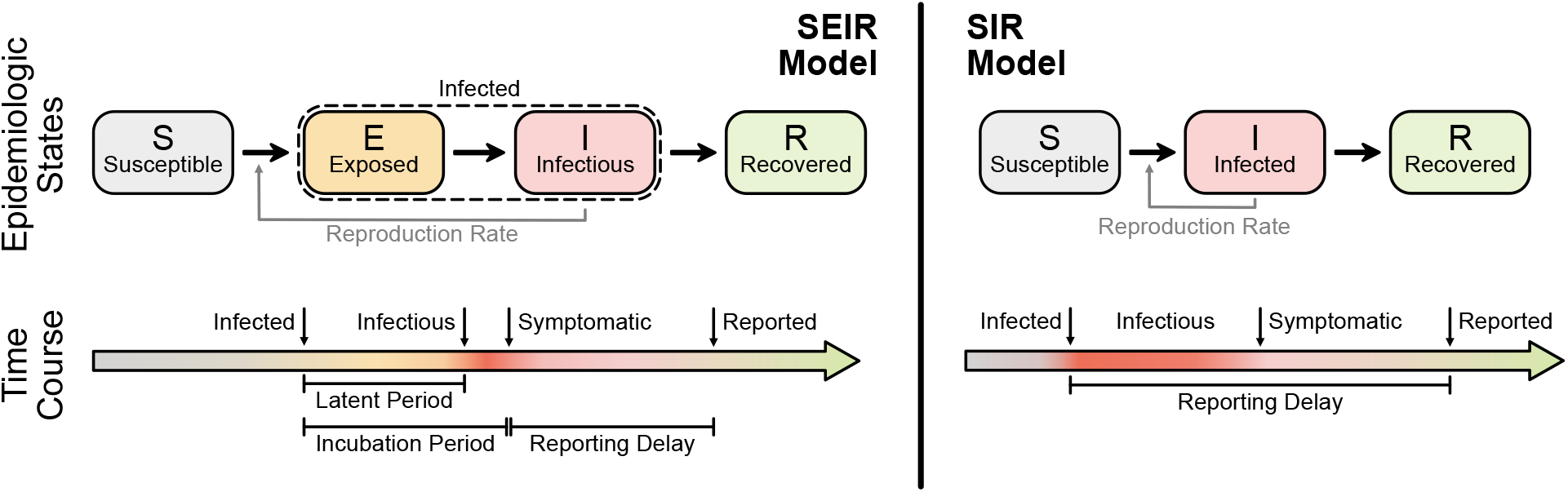
Illustration of two basic compartmental models in epidemiology. The SEIR model (left) captures the basic steps that infections passes through: A healthy person becomes infected but not infectious (leaves S, enters E); after some time (’latent period’) the person becomes infectious (leaves E, enters I) but symptoms only show after some incubation period; after some time the person is no longer infectious (leaves I, enters R), which can have several reasons including isolation, conventional recovery, or death. The SIR model (right) is the most basic compartmental model and does not distinguish between infectious and infected: A healthy person becomes infected (leaves S, enters I), immediately begins to infect other persons but only shows symptoms with a delay. After some time the person “recovers” (leaves I, enters R), which again includes isolation, recovery, or death.

This assumes a spreading rate *λ* for infected people to infect susceptible people (who they meet randomly) and a recovery rate *µ* for infected people to recover. These differential equations can be extended to include various different compartments, in order to better resolve the temporal course of the disease, but typically keep the mean-field assumption of a well-mixed population unless evaluated on some (typically unknown) network. In this case, additional compartments reflect spatial information.

Observed case numbers are always delayed from the true infection date (Fig. 2). In general, when a person becomes infected, the onset of symptoms is delayed by the incubation period. Upon symptom onset, it typically takes a few days until the person undergoes a test and the case is reported (although some people are tested before symptom onset, e.g. if contacts are traced or tests are performed at random “Stichprobe”). However, for the modeling, one is usually interested in the actual time when a person becomes infected — but this information is not directly available in real-world data. One either works with the reporting date or with the dates of the symptom onset (epi curve) that can be reconstructed e.g. via questionnaires and imputation methods. Note that even symptom onset dates are still delayed with respect to the true infection dates due to the incubation period. For the reporting dates a second delay occurs between symptom onset and report, unless an asymptomatic case is discovered in random testing. For the example models in the following, we synthetically generate observed cases — symptomatic or reported — by convolving the infected cases with a distribution of incubation periods or reporting delays, respectively (Fig. 2).

**FIG. 2.**
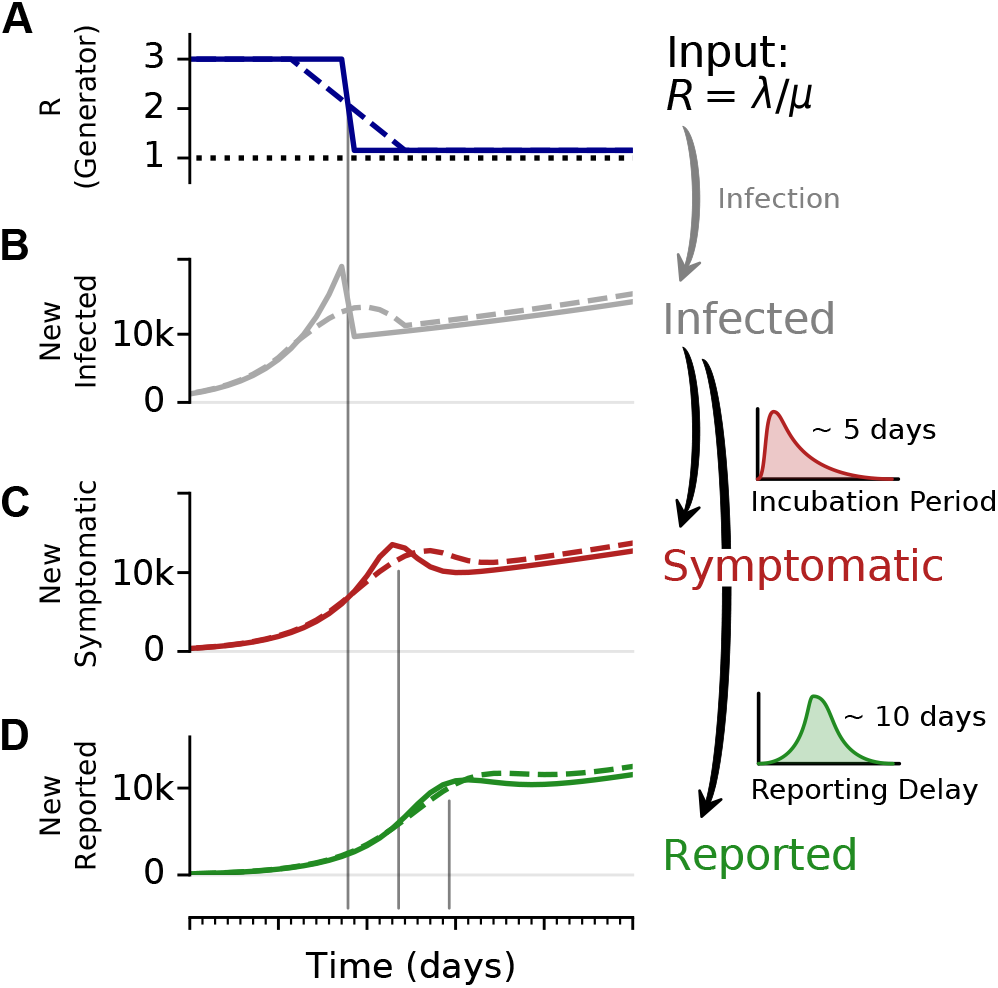
A change-point in *R* can lead to a transient decrease in case numbers. To illustrate the effect of a change point, and the delays in observing symptomatic and reported cases, we consider an SIR model with a fast or slow decrease of R, and generate synthetic case numbers. A: The reproductive number *R* exhibits a change point from *R* = 3 to *R* = 1.15, with a duration of either 1 day (solid) or 9 days (dashed). B: The number of new infections can show a transient decrease caused by the change point in *R*, even though the underlying dynamics are always in the exponentially growing regime of *R* > 1. Such a decrease can be misinterpreted as *R* < 1. The number of C new symptomatic cases, and D reported cases is generated by convolving the new infected with a log-normal incubation period (median 5 days) or reporting delay (median 10 days), respectively. Note that the convolution shifts and smooths the curve of the new infected. Nonetheless, the counter-intuitive effects of a transient decrease in case numbers caused by a change point, is still very well visible (See Fig. 4 for the challenges in estimating *R* around the change point.)

### B. Model-free estimation of reproduction number *R_t_*

#### Definition of *R*

The reproductive number *R* quantifies how many susceptible persons are on average infected by one infected person. If one infected person infects on average more than one other person *(R* > 1), then case numbers are growing exponentially. In contrast, if less than one other person gets infected *(R* < 1), then case numbers are declining. Therefore, *R* = 1 marks the critical transition between growth and decline of case numbers. Last, note that *R ≈* 1 means that new infections keep occuring at their current levels (which may be high, depending on when and how *R ≈* 1 was reached).

Estimating the reproductive number *R* in principle can be done in two manners, either by inferring it from observed case numbers, or by following infection chains step by step (which is not discussed here). If one infers it from observed case numbers, there are a number of possible approaches. Some approaches are summarized in Fig. 4 and detailed below. All of these approaches can be applied to the epi curve (day of symptom onset) or to the reported cases (day or reporting). In the following, we assume that they are applied to the epi curve.

The most straight-forward definition of the reproductive number assumes a reproductive process with offspring generation, such as a branching process [2]. For this, one assumes a generation time *g* in which an infectious person can generate offspring infections. In the simplest case, one could consider that offspring infections occur exactly after one generation time *g*. This allows to infer the reproductive number *R* precisely:

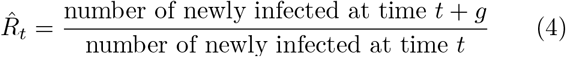

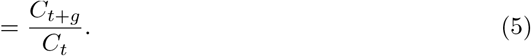

In reality, these newly infected case numbers *C_t_* have to be approximated, e.g., by using new symptomatic cases or new reported cases. Moreover, the generation times *g* of each infection are widely distributed, so that using the average value *g* (or an estimate of it) is used as a further approximation.

When going into detail, there are two different conventions for the timing of the estimated reproductive number 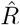 with respect to the case numbers *C_t_* (Fig. 3). Above, we consider 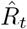 to characterize the number of future infections that are caused by infections at time *t* (left-edge convention). Alternatively, one can consider 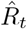 to characterize the number of infections at time *t* that were caused by the past pool of infected (right-edge convention), defined as

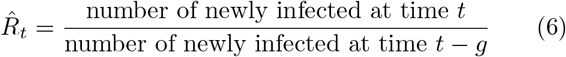

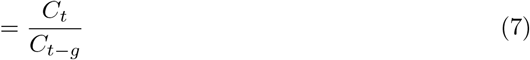

**FIG. 3.**
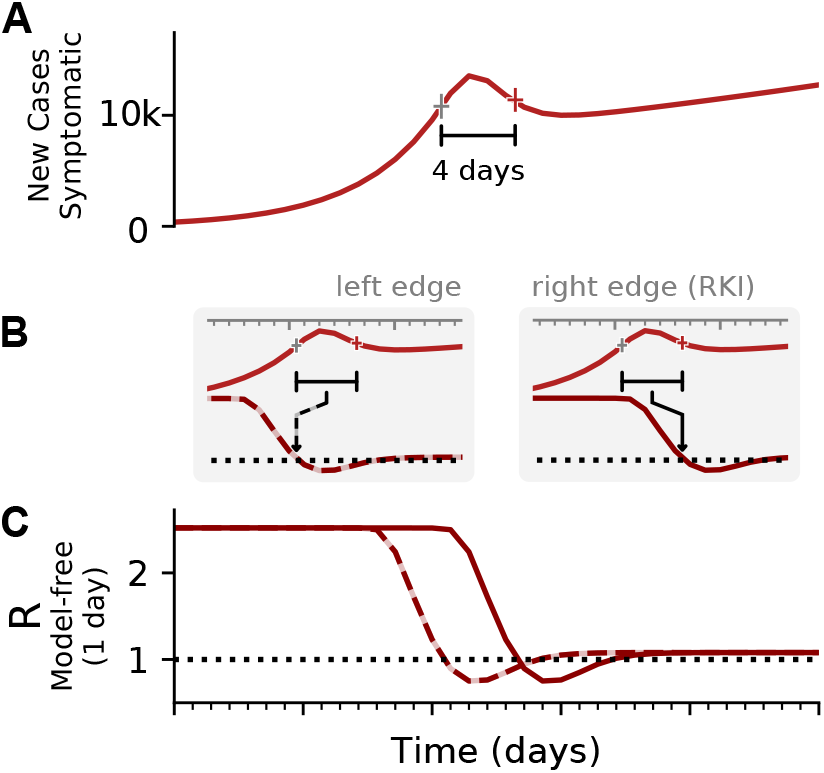
Two different conventions to define the reproductive number *R*: Infections in the future or infections from the past. **A:** Synthetic data for new symptomatic cases. The marked interval indicates an assumed generation time of 4 days. **B:** The basic reproductive number can be defined either on the left edge of the generation interval (left, dashed line), describing the average number of future infections that are cause at time *t*, or on the right edge of the interval (right, solid line), describing the average number of infections at time *t* that were caused by the past ones. **C:** Depending on the convention, the resulting curve of *R* is shifted by the generation time *g*. Note that in both cases the *R* is estimated erroneously to fall below *R* = 1, although in the underlying model it was was *R* > 1 all the time. This is an effect of the SIR dynamics together with a change point in the underlying *R*. (See Fig. 4 for model details, and Figs. –for other parameters).

The results for 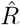 are exactly equivalent, apart from a shift in time by exactly *g*. However, the distinction between left-edge and right-edge convention and the associated time-shift crucially matter when discussing changes in *R_t_* with respect to interventions.

#### 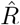 as calculated by the RKI

Real-world data are often noisy, and therefore averaging over a certain time window can help to smooth the estimate. This procedure is used in two variants by the RKI, smoothing over four days or over seven days. The details of the procedure are documented in detail in Ref. [3]. For both smoothing lengths, they assume a constant serial interval (generation time) of *g* = 4 days (Fig. 4) and the right-edge convention. The four-day smoothing has the advantage that it reacts a bit faster, the seven-day smoothing has the advantage that it smooths better the relatively strong variations. In particular,

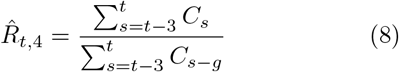

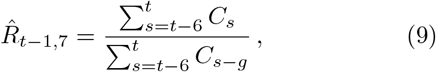

**FIG. 4.**
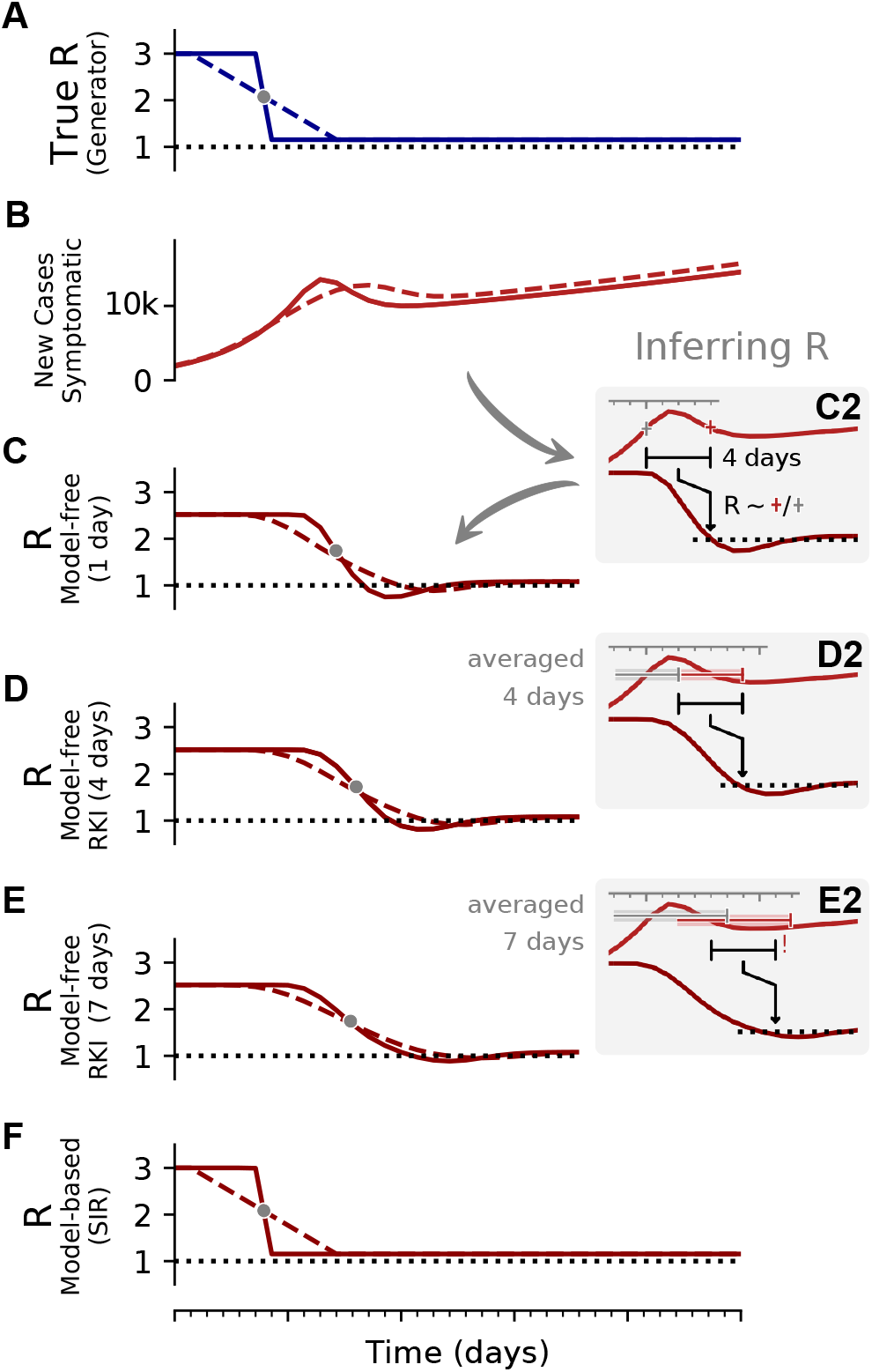
The inferred reproductive number 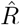 depends on the inference method. **A, B:** Synthetic data for new symptomatic cases generated with SIR dynamics from an underlying *R* with one change point of duration 1 day (solid) or 9 days (dashed). **C:** Model-free inference of 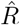 based on the ratio of case numbers at time *t* and time *t−g*, marked by a red and gray cross (inset), respectively (’right-edge convention’, cf. Fig. 3). **D:** Model-free inference of 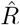 following the Robert Koch Institute convention, i.e. using the definition of C but with averaging over a window of the past 4 days (inset, red and gray bars). **E:** Same as D but averaging over 7 days. Note the overlap of intervals. All the model-free methods (C–E) can show an erroneous estimate of *R* < 1 transiently, due to the change point in the underlying true *R*. **F:** The inferred 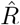 using change-point detection with an underlying dynamic model (SIR) does *not* show a transient erroneous *R* < 1 period. If the underlying dynamic model corresponds well enough to the true disease dynamics, then this approach reproduces the true *R* that was used to generate the data.

where *g* = 4 is the assumed generation duration, and the averaging is done over 4 and 7 days, respectively. Note the shift by one day in the 7-day version Eq. (9).

#### Model-free methods also build on assumptions

Clearly, when using *model-based* methods, assumptions go into the model itself; but also when using what we call *model-free* methods, assumptions have to be made. In particular, the core assumptions behind the model-free approach to estimate *R* that we discussed so far are that every new infected person infects on average *R* persons, and it does so precisely *g* days after becoming infected. As is the case in modelling, these assumptions present a simplification of the complex real-world dynamics. Whether a chosen way to answer a given question is reasonable or not depends on the specific question one asks (every question may need its own model simplifications and type of data set), on the quality of the data, and on how well the *relevant* real-world dynamics for the question are captured in the simplified model. For the question of whether case numbers are increasing or decreasing *in general*, the above method of calculating *R* has proven very useful.

### C. Model-free methods versus model-based methods to infer the reproductive number

In order to demonstrate potential issues when inferring the reproductive number *R*, we systematically compare the model-free methods with model-based methods (akin to our analysis of *λ^∗^* in [1]) on synthetic data from an SIR model (Fig. 2). With model-free methods, we refer to inference methods for *R*, which do not explicitly incorporate disease dynamics (SIR). The three methods we presented above belong to this group. These methods to estimate *R* are straight forward and easy to implement. However, they might lead to biased estimates when *R* is changing rapidly. More precisely, in the following we show that these methods (1) smooth out fast changes in *R*, (2) produce some delay compared to the underlying *R*, (3) the estimate depends on the assumed generation time, and (4) around change points they may return transiently *R* < 1, even if the true value was never smaller than 1. While these methods have the above limitations when *R* is changing quickly, they are still very useful for an easy-to-obtain estimate of *R*.

#### 1. Model-free methods may smooth out fast changes

In Fig. 4, the 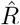 that is inferred by model-free methods undergoes a smoother change than the true *R*. The smoothing has two origins: First, when using the sliding-window of four or seven days (RKI methods), multiple days are combined to obtain an 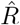 alue for one day. Second, 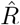 has to be calculated from the daily new symptomatic or reported cases (Fig. 2 C, D), because the dates of infection (Fig. 2 B) are not directly accessible in real-world data. As discussed before, symptom onset and reporting date are delayed from the infection date. Because the delays vary from case-to-case, these two curves are smoothed out compared to the infection curve (in other words, the smoothing originates from the variance in incubation period and reporting delay, see later Fig. 12 in the section about testing). Hence, if smoothing is not explicitly incorporated in the inference of *R*, fast changes appear slower than they truly are, and successive fast changes may appear as a long transient.

#### 2. Model-free methods produce delayed estimates that are difficult to interpret

In our example in Fig. 4, we estimated 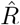 based on the number of new symptomatic cases as produced by our synthetic disease model. The 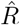 of all three model-free methods is shifted in time compared to the true *R* (Fig. 4 A).

How does one interpret the shift and where does it come from? To interpret the shift and compare between the different methods, we focus on the time point where half of the steep step in *R* has been detected (gray dots). This shift has multiple contributions. One contribution originates from using the dates of symptom onset, which is shifted on average by the incubation period (in our example *≈*5 days). This generates the 4–5 day shift of the one-day method (Fig. 4 C). Because the incubation period is not constant and typically asymmetric, there is an additional asymmetric distortion towards either direction, depending on the shape of the actual distribution of incubation periods. Another source for the shift comes from the time average, which explains the additional (approximate) 1–2 day shift in the four-day and seven-day methods employed by the RKI (Fig. 4 D, E). Because of the specific definition of the position of the 4 and 7-day window of the RKI (see eq. 8, 9), the two versions of 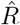 have a very similar average delay of 5–6 days in total with respect to the true *R*.

Both, the variable incubation time and the time averaging also impact the start- and end-points of the change in a non-trivial manner. In combination, multiple sources cause shifts that can point into opposite directions. While the sources can be identified conceptually, the combined effect cannot be perfectly disentangled or compensated.

Due to multiple sources of shifts and smoothing, a simple post-hoc shift of the 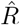-curve cannot reproduce the true *R* around a change point. For example, a shift of Fig. 4D by 5 days would suggest a start of the change point before it starts in reality (Fig. 4 A). This fact has led to multiple prominent misunderstandings in relation to the RKI data and the effects of governmental interventions. Instead of shifting curves to partially correct for one or another potential delay, an inference of *R* using model-based methods can account for this and other potential biases. When using a good model, such a model-based approach returns the correct *R* with the correct steepness and time point (Fig. 4 E, for technical details, see Methods in [1]).

#### 3. R-estimates depend on the assumed generation time

The assumed generation time *g* impacts the magnitude of the estimated reproductive number 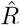 (Fig. 5). We exemplify this effect using the method of the RKI (4-day average), where we vary the assumed (constant) generation time *g*. In particular, the chosen generation time *(g* = 2, 4 or 8) affects the initial plateau (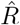‘ 1.6, 2.5 and 6.4 respectively), the duration of the inferred change, and the depth of the transient underestimation. This small example shows that estimating the magnitude of the reproductive number from observed case numbers without knowing the precise generation time can be challenging.

**FIG. 5.**
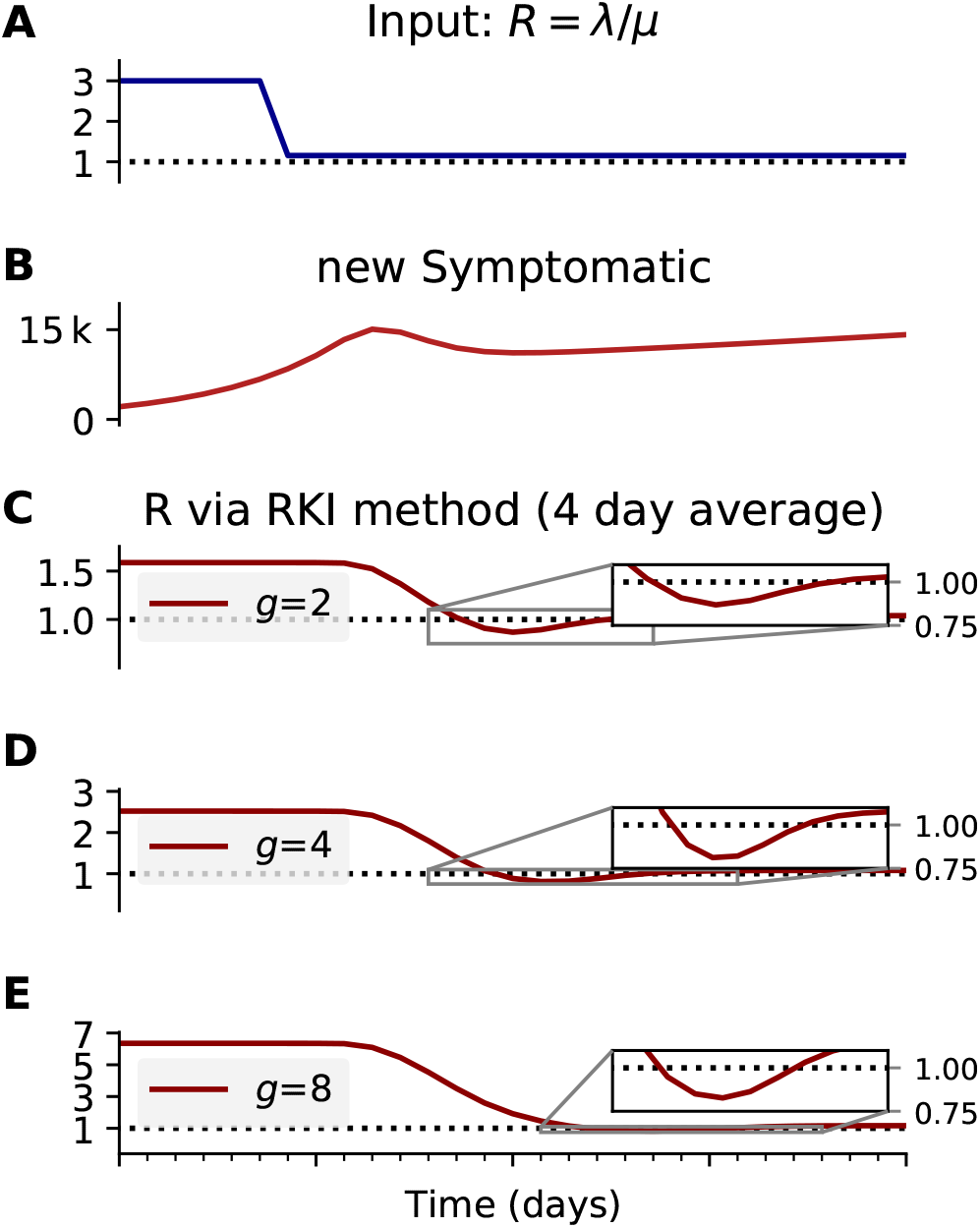
The inferred reproductive number depends on the assumed generation time g. **A, B:** We generate synthetic data using SIR dynamics with time-dependent *R* including a 1-day change point (A) that yields new symptomatic cases with transient decrease (B) despite all *R* > 1. **C–E:** Using the RKI convention to infer 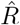 (4-day average, right-edge convention), we demonstrate how generation times *g* result in different 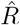 curves. In particular, we find different initial levels of *R* (left plateau), differently long crossover duration (time from left plateau to right plateau), and differently deep transients of *R* < 1 (insets).

#### 4. *Model-free methods may return erroneous transient periods of R* < 1 *at change points*

In our examples (Figs. 4 and 5), we consider that *R* changes rapidly from *R*_0_ = 3 to *R*_1_ = 1.15 within one day (full lines). Such a sudden change leads to a transient decrease in new case numbers — despite *R* being always *>* 1. How can there be decrease in new cases although *R* > 1? The transient decrease results from the pool of infected suddenly infecting considerably less people. This decrease in infections causes the sharp peak and a sudden drop in new infections (Fig. 2 B, solid line). It then carries over to the number of new symptomatic and new reported cases, with the respective delay and smoothing (Fig. 2 C, D]). This transient decrease depends on the duration of the change point: While it is strongest for steep changes, it also occurs for a change point with a transient time of nine days (Fig. 2, dashed line).

Naively, a transient decrease might be interpreted as a transient *R* < 1, but that is not the case here. A model-free method cannot distinguish between different causes for transient decreases in case numbers, being it due to transient non-linear effects (Fig. 2) or due to a true exponential decay *(R* < 1). The model-free methods in our example (Figs. 4 and 5) correspondingly yield non-negligible periods of *R* < 1, even though the underlying model dynamics have *R* > 1 always. Model-based approaches, on the other hand, can account for transient non-linear effects if included in the model, e.g., as change points, and — if the model is correct — even reproduce the true underlying dynamics (Fig. 4 F). To conclude, if one infers *R* in a model-free manner, by computing ratios of case numbers, one interprets reductions in case numbers as 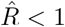 (Fig. 4 C–E). After strong decreases of the true *R* this may be an incorrect interpretation.

#### 5. Well chosen model-based methods can reconstruct complex disease dynamics

When the chosen model describes the true disease dynamics well, robust inference of the true underlying reproduction number (and other parameters) is possible. To demonstrate the robustness of model-based inference, we generate synthetic data using an SIR-model as inferred from case numbers in Germany between March 2 and April 21 [1] (Fig. 6). The Bayesian model inference can recover the reproductive rate (Fig. 6 D, F), whereas with the model-free method, the recovered *R* is slightly biased (Fig. 6 C). Note, however, that the chosen model has to match at least approximately the disease dynamics, to allow a good inference. This is why we used different models to assess the robustness of our results in Ref. [1] (SIR: Fig. 3, SEIR-like: Fig. S3, SIR without weekend modulation: Fig. S4).

**FIG. 6.**
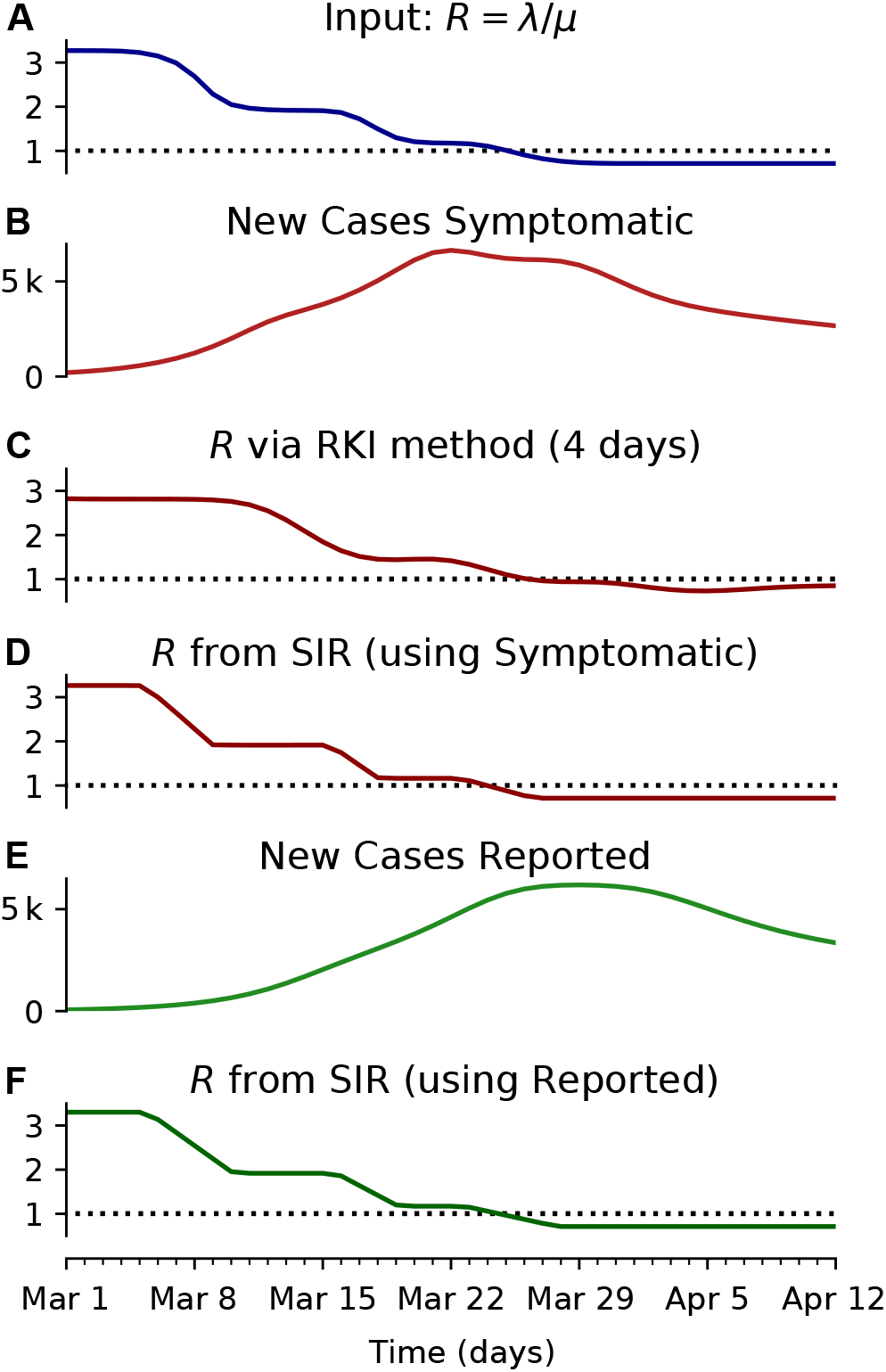
The model-based methodology yields consistent results irrespective of whether it is applied to the new reported cases or the new symptomatic cases. (e.g. obtained by nowcasting). **A:** Time-dependent reproductive number as inferred from case numbers in Germany [1]. **B:** Synthetic data for new symptomatic cases generated with SIR dynamics from the underlying time-dependent *R*. **C:** Inferred 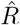 from new symptomatic cases using RKI method (4 days generation time, right-edge convention) would reproduce step-like behavior (no noise present) but drops below *R* = 1 (dotted line) already after the second change point (note that curve is shifted and smoothed compared to input *R*, cf. Fig. 4). **D:** Inferred 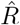 from new symptomatic cases using change-point detection with dynamic model (SIR) correctly reproduces the input. **E:** Synthetic data for new reported cases generate with SIR dynamics as in B (cf. Fig. 2). **F:** Inferred 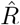 from new reported cases (E) using change-point detection with dynamic model (SIR) also correctly reproduces the input. Note that both, D and F show sharper steps because of the assumed piece-wise linear change points in the model, and that they perform so well because they employ the true dynamic model that is used for the synthetic data. Both are model assumptions that need to be justified in our approach.

## III. WHAT CONCLUSIONS CAN ONE DRAW FROM A BAYESIAN ANALYSIS?

### A. Modeling background

When the Coronavirus-pandemic arrived in Germany we set out to model the spread of the disease as rapidly as possible. Thus, our model from the start was aimed at giving estimates with their corresponding error bounds based on the data available at that time. To this end we decided to use a Bayesian strategy as it allowed formulating well-documented assumptions on those aspects not available from data at that time. Within the Bayesian framework these assumptions can and should be replaced by data as soon as these become available, and we implement such an improvement below for the case of information on symptom onset times that have become available in the meantime. Given such new data it will also be interesting to evaluate post-hoc the assumptions and the performance of our model. This will also give some guidance as to whether to employ a model of this kind again in a new scenario (another disease outbreak or pandemic) where some relevant data will also not be available immediately. We note that taking these steps is the intended development in Bayesian inference.

We also note that all statistical procedures come with their own assumptions, e.g. on distribution of the data, models of measurements and random errors. Bayesian analysis is no exception to this rule; in our view the only difference is that modeling assumptions are not taken for granted based on the long-established used of a method (say, a t-test) but need to be formulated anew for each case. The fact that the assumptions are hand-tailored to the application case may seem subjective sometimes; yet, similar assumptions are being made, more tacitly perhaps, in other frameworks, as well. This said, it is nevertheless important to question and discuss (our) modeling assumptions and to test the sensitivity of our results to these modeling assumptions. We have already concisely discussed our assumptions in the main manuscript [1], but we here give a much deeper, broader and more educational treatment.

### B. Bayesian inference as reasoning under uncertainty, bound to be updated

The results of a Bayesian analysis at some publication time point *T* represent what we should consider most plausible at that time point *T*, given the knowledge available at *T* (causes and data known at *T)*. These results represent something that we should be able to agree on given the knowledge at *T* (and some practical constraints, see below), but these results may change given more information at a later time *T* +∆*T*. Changing one’s mind with the availability of additional information is designed into Bayesian inference as “the logic of science” (E.T. Jaynes,[4]) from the start. In other words, scientific inference and the associated models are bound to be updated. The important question is thus not whether a model is correct in absolute terms, but whether it was possible to agree on the model (and the inference provided by it) at time *T*, and also if the inference provided at *T* was robust, for example in the sense that the credible intervals for the model parameters at *T* comprise those obtained at *T* + ∆*T*.

From this perspective, it is obvious that now, more than a month after finalization of our published analyses on April 21, new data have become available and that the model can, and should, be improved accordingly. Important data in this respect are data on reconstructed infection dates which at present take about 7 days to come in for at last 80% of the cases (Fig. 12), and took even longer during the early stages of the outbreak. We present results obtained using these data below and compare them to our published results.

### C. Conditions for plausible alternative models entering model comparison

A frequent, and important misunderstanding around Bayesian model comparison is that one is allowed to formulate very many models at random and then let the data decide on the best model via the Bayesian model evidence (or the LOO-scores). This notion fails to notice that the model evidence *p(D|M_i_)* is only one part of the decision on the preferred model. The formal equation for deciding between models *i* and *j* would be:

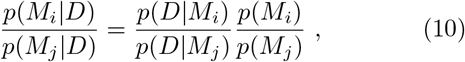

i.e. taking such a decision entails accounting for a-priori plausibility of different models, i.e. *p(M_i_)* and *p(M_j_)*. While it is customary to assign equal a-priori plausibility to all the models being considered, this does not mean that just any model qualifies for use in this decision procedure. Rather, each model subjected to a model comparison needs to be well justified. This is one of the reasons why we did not consider for example models of sustained, constant drifts in the effective spreading rate *λ^∗^* (or, equivalently the reproductive number R), as we did not come up with plausible explanations for such a behavior (except perhaps arguments based on herd-immunity, which seem implausible now, in the light of second waves of infections and a recent rise in *λ^∗^* from its all-time low, and also in the light of country to country comparisons, Fig. 7).

**FIG. 7.**
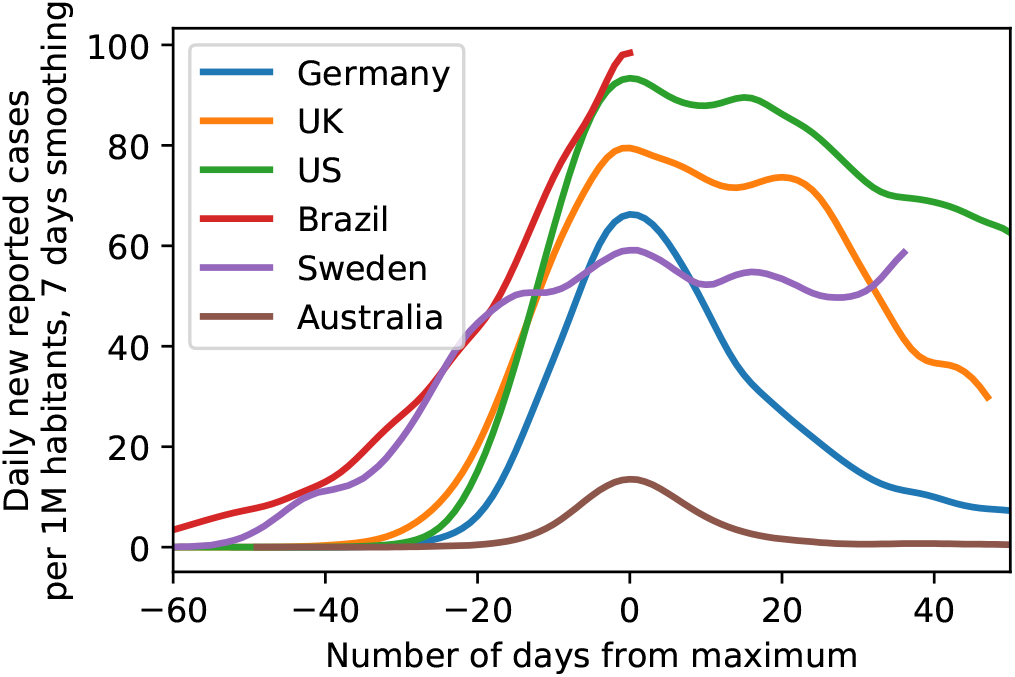
Comparison of the case numbers per one million inhabitants of exemplary countries as illustration of the range of possible case numbers developments. Note how both the peak height as well as peak width of some countries are considerably larger than for Germany, providing evidence against saturation effects (’herd immunity’) in Germany (Data until June 3, 2020).

On a practical note, useful modeling also has to reflect certain limits on model complexity in relation to the available data, and also computational resources. Known phenomena, that can nevertheless not be modeled must therefore often be integrated into noise terms that are designed accordingly (as was done with the modeling of observation noise in our case, instead of using full stochastic differential equations). The best that can be done then is to investigate the sensitivity of results with respect to the simplifying assumptions that have been made.

It is also in order to explain in simple terms how results of a Bayesian analysis may be interpreted: In the Bayesian framework probabilities are measures of the plausibility of statements about the world, given our present knowledge (see [4] for the exact mathematical derivation of this statement). Thus, the results of Bayesian parameter inference indicate credible (plausible) ranges in which we should assume the unknown parameters to be. Assuming them to be elsewhere with high probability would be inconsistent with the information we have. In this sense, these credible intervals may form the basis for decisions we have to take.

### D. Models as competing causal explanations of data

Last, we note that the notion of causality resides only in the construction of the models — with different models incorporating different possible causal explanations of the data (e.g. in the form of differential equations for the disease dynamics). Performing model comparisons then selects more plausible over less plausible explanations but does not provide a proof of causality in the strict sense advocated for example by Pearl [5] or by Ay and Polani [6]. Yet, fulfilling the formal criteria for causality in this strict sense would need multiple replications of the pandemic process, each time with different settings of the relevant variables, such as interventions. Even when treating the SARS-CoV-2 outbreaks in different countries around the world, with their different interventions (or lack thereof), as replications establishiung formal casuality may remain an elusive goal due to multiple other variations from country to country. In sum, the results of our Bayesian analysis must be seen as a search for the most plausible causal model of the data, given the data available at the time of analysis, and as providing credible ranges of the parameter values relative to this most plausible model.

Later, discussions (such as the one presented here) of the selected models and the inferred parameter ranges should then investigate and update modeling assumptions, and reason whether the causal model can be maintained, or not.

When analyzing improved data that reflect the dates of symptom onset rather than case reports to improve our modeling, we find that both the preference for a three change-point model as well as the inferred parameter ranges do not change drastically, and we maintain our original interpretation of the pandemic process and the effectiveness of governmental interventions.

Last, alternative models assuming herd immunity as a reason for the sustained observed drop in infection rates still do not seem plausible to us in the light of rapidly surging second waves or sustained high levels of new infections (such as in Sweden, see Figure 7).

## IV. MODEL EVOLUTION

Modeling efforts at the beginning of an epidemic outbreak are aimed at providing a rough but timely and robust description of the disease outbreak, making use of data that is available (and easily accessible) at that time (Tab. I). Later modeling efforts, in contrast, can make use of more detailed data and provide deeper insights into how the outbreak unfolded. While these latter models are useful for a better understanding after the fact, they cannot be applied early on due to a lack of data, and often cannot inform decisions sufficiently fast. However, a comparison of early and later models can provide important insights about the robustness and usefulness of the early models with respect to the later ones (here usefulness means that the early models describe the epidemiological parameters and their uncertainties well enough to inform decisions).

**TABLE I.** Data sources differ in availability, the detail they provide, and accessibility. For our previous study [1], modelling needed to be fast; we used the JHU data from GitHub because it was available early, it is easy to access (machine readable) and it is the unofficial go-to resource on case numbers. Note that some sources (red cross) need manual extraction of the data from a plot — a process that, even when assisted [14], introduces uncertainties. Also note that only some of the listed sources are also accessible in their past, as-was state (for instance, the dashboards only display the most recent data, in real-time).[7][8][9][10][11][12][13]

| available since | source | machine readable | manual extraction needed | reported cases | known symptom onsets | imputed symptom onsets | nowcasting | notes |
| --- | --- | --- | --- | --- | --- | --- | --- | --- |
| Jan 22 [7] | JHU dashboard (ArcGIS) |  | x | x |  |  |  |  |
| Feb 2 [8] | JHU dashboard (GitHub) | x |  | x |  |  |  | first commit with case numbers on Feb 4 |
| Mar 4 [9] | RKI situation report (pdf) |  | x | x | x |  |  |  |
| Mar 20 [10] | RKI dashboard (ArcGIS) |  | x | x |  |  |  | known symptom onsets added ~2nd half of April |
| Apr 6 [11] | RKI dashboard (API, csv) | x |  | x | x |  |  |  |
| Apr 9 [12] | RKI Epi Bulletin (pdf) | | x | x | x | x | x | nowcasting introduced in bulletin 17/20v1, includes $R_t$ estimate, note the transient $R < 0$ |
| Apr 15 | RKI situation report (pdf) |  | x | x | x | x | x | nowcasting initially only in German |
| May 11 [13] | RKI resources on nowcasting (xlsx) | x | | x | x | x | x | includes $R_t$ estimate |

For the case of the COVID-19 outbreak in Germany, the initially available data were sorted based on date of reporting, where the reporting occurred after an unknown delay between symptom onset and report. Only later, data organized by time of symptom onset, the so-called epi curve, became available. Even after their initial release, these data were still updated and refined (see Fig. 10); also note that data for symptom onsets still take some time to arrive and be compiled, i.e. the delay between symptom onset and testing/reporting is still considerable (see Tab. I). In particular, this means that *reliable* epi curve data for the date of April 21, our analysis cutoff date in [1], were not available on that day but only considerably later (cf. Tab. I). Now that these data are available, however, we can compare models based on data organized by reporting date (modeling the reporting delay and incubation period) with models based on the epi curve (modeling the incubation period, only).

### A. Model updates based on time of symptom onset and comparison to previous results based on time of reporting

Ideally, modeling of an epidemic outbreak should rely on data organized by infection date — yet, such data are rarely available outside of the analysis of individual, well-confined infection chains. The next best option are data organized by date of symptom onset — the epi curve. Normally, symptom onset precedes the test and report in time. Thus, the epi curve is only available after a certain delay, which can be substantial. Furthermore, the time of symptom onset may remain unknown for a significant fraction of reported cases. If so, then reconstructing the epi curve requires data imputation and further modeling (e.g. nowcasting [18, 19]), which may further delay the availability of this curve and introduce additional sources of uncertainty. At the initial stages of an outbreak, one may therefore decide to analyze data organized by reporting data, and to model the relevant delays.

For a comparison of analyses, it is important to understand how the curve of reporting dates and the epi curve are linked. Both curves originate from the curve of initial infections by a convolution (see again Fig. 2). The epi curve is the curve of initial infections convolved by the distribution of incubation periods, while the curve based on reporting date is the curve of true infections convolved by the (less well known) distribution of delays between infection data and reporting date. Technically, a report can also happen before symptom onset, albeit this is typically rare^1^. Therefore, the curve of reporting dates is not exactly a convolution of the epi curve with an additional delay distribution.

We have reanalyzed the initial stages of the outbreak until April 21 based on the epi curve that has become available (cf. Tab. I). In Figure 8, we show a comparison of our analysis where the SIR model with three change-points is applied to RKI data, using either the reporting date or the date of symptom onset (for the SEIR model, cf. Figs. 17 and 19 at the end of the document).

**FIG. 8.**
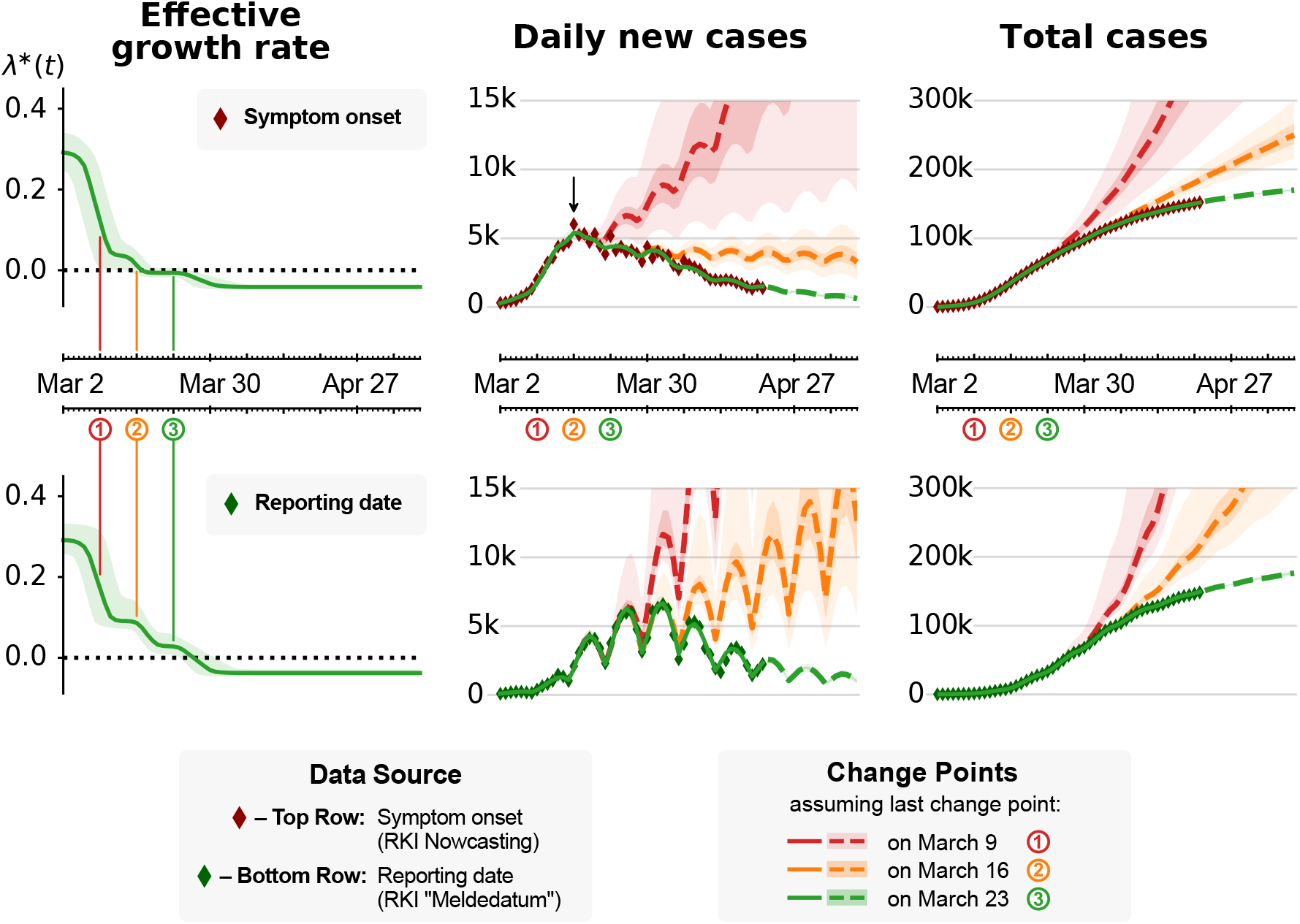
Model-based inference is consistent when based on symptom onset (top) or reporting date (bottom). We repeated our SIR-model based inference (Fig. 3 in [1]) that used JHU data [15], now using the date of symptom onset (red diamonds, top) [16] and the reporting date (green diamonds, bottom) of daily new cases as reported by the RKI [17]. **Note:** We currently do not incorporate the uncertainties that are introduced by nowcasting (red diamonds, top), compared to using the reported cases. This leads to *over-confident* parameter estimates, including the effective spreading rate *λ^∗^(t)*; the shown uncertainties are underestimated. **Left:** Effective growth rate *λ^∗^(t)* inferred by the model. Dates of the three main public interventions are indicated by colored circles and vertical lines. The values of *λ^∗^(t)* before and after *all change points* is consistent across both data sources. Note that, when the symptom onset is used, *λ^∗^(t)* drops to zero already after the second change point. Still, only after the third change point *λ^∗^(t)* becomes sufficiently negative to cause *decreasing* daily new case numbers. **Center:** Daily new case numbers. Dashed lines show inferred case numbers assuming that the last two (red) or the last one (orange) change points were excluded. The weekday-dependence in daily new reported cases is already accounted for when using symptom onsets (top). **Center, Top:** Although *λ^∗^(t)* already dropped to (slightly-below) zero as of the second change point, daily new cases do not decrease if the third change point is excluded (orange). Note the arrow: Due to the transient decrease in new cases after change points (cf. Fig. 4) as well as the delay between symptom onset and reporting (cf. Fig. 4), the peak that corresponds to maximum daily new infections is located already around March 16 (for symptom onsets); yet note again that this does *not* mean that new cases would have declined rapidly already after the second change point (see the orange curve). **Right:** Total, cumulative case numbers.

These complementary results do not change our main inference result presented in [1]. Specifically, model comparison still favors the three-change-point models over their simpler counterparts (Tab. II), and only the third change point leads to a value of the effective growth rate *λ^∗^* that is clearly below zero. Importantly, the growth rate has to be sufficiently below zero to cause a notable decrease in new infections. At the quantitative level, however, the model based on the epi curve (Fig. 8, top row) suggests a larger drop introduced by the first change point compared to the model based on the reporting dates (Fig. 8, bottom row), and consequently smaller drops induced by the second and third change point. This effect is more pronounced in the SEIR model (see Fig. 19 at the end of the document). These quantitative changes are driven by the epi curve dropping faster than the curve of reporting date (Fig. 8, middle column). Note, however, that we did not yet include in our analysis the uncertainty of the epi curve from the nowcast data imputation, nor did we consider the effects of potentially missing data at the time of analysis. For example, comparing the maximum of the epi curve in mid March, using either data available mid April or end of May still shows clear differences (Fig. 10. This shows that the case number counts of the epi curve and the associated nowcast are being updated for several weeks after the putative symptom onset date.

In sum, we conclude that the original model based on data organized by reporting date was useful to understand disease dynamics in the absence of the epi curve and robust in the sense that its main results still hold.

**TABLE II.** Model comparison: Using leave-one-out (LOO) cross-validation, we compare the SIR and SEIR model variants using the epi curve as data (Figs. 17 and 19). Lower LOO-scores represent a better match between model and data (pLOO is the effective number of parameters).

| Model | # c-pts. | LOO-score | pLOO |
| --- | --- | --- | --- |
| SIR main | 0 | $900 \pm 13$ | 6.36 |
| SIR main | 1 | $774 \pm 14$ | 12.72 |
| SIR main | 2 | $755 \pm 13$ | 12.17 |
| SIR main | 3 | $725 \pm 15$ | 19.66 |
| SEIR-like | 0 | $900 \pm 14$ | 6.65 |
| SEIR-like | 1 | $749 \pm 12$ | 8.05 |
| SEIR-like | 2 | $739 \pm 13$ | 10.28 |
| SEIR-like | 3 | $726 \pm 14$ | 14.04 |

### B. At most minor differences between results based on RKI versus JHU data sources

At the beginning of the outbreak, data were made available on a daily basis both by Johns Hopkins University (JHU) and the German Robert Koch Institute (RKI). Both sources initially provided only reported cases (in text form), with the JHU resources providing data faster and with a better interface for automated analyses. The RKI resources were updated only a few days later, as information has to be transmitted from regional agencies to the RKI, whereas the JHU data for Germany are gathered from a few reputed online media (Berliner Morgenpost, Taggesspiegel and Zeit Online [20]). However the JHU resources have been partially criticised for lacking quality control (see issues section on the Github page [15]). We therefore compared the JHU data used in [1] to the official RKI count (Fig. 9) and have rerun the analysis using the RKI reported cases (the “Meldedatum”, Fig. 15 and 16). The differences are clearly very minor.

**FIG. 9.**
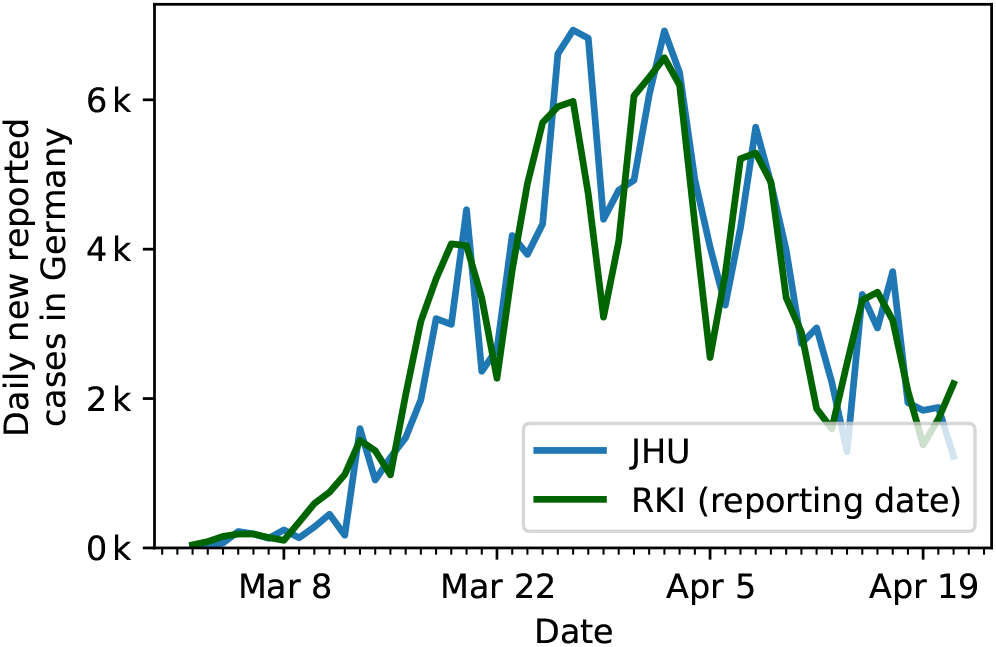
Comparison of the German case numbers as published by the Johns Hopkins University (JHU) used in our previous publication [1], to the case number of the Robert Koch Institute (RKI).

## V. IMPACT OF TESTING

Our modeling depends on reported case numbers, which in turn depend on testing. Throughout the COVID-19 spread, test availability, test requirements and known case numbers changed continuously over time (Fig. 10). Such an inconsistent and fluctuating data-acquisition obviously introduces additional sources of uncertainty. While we decided to exclude the effects of testing in previous models, concerns about results derived from data that stem from inconsistent testing should be taken seriously. Thus, we analyze possible distortions in more detail.

**FIG. 10.**
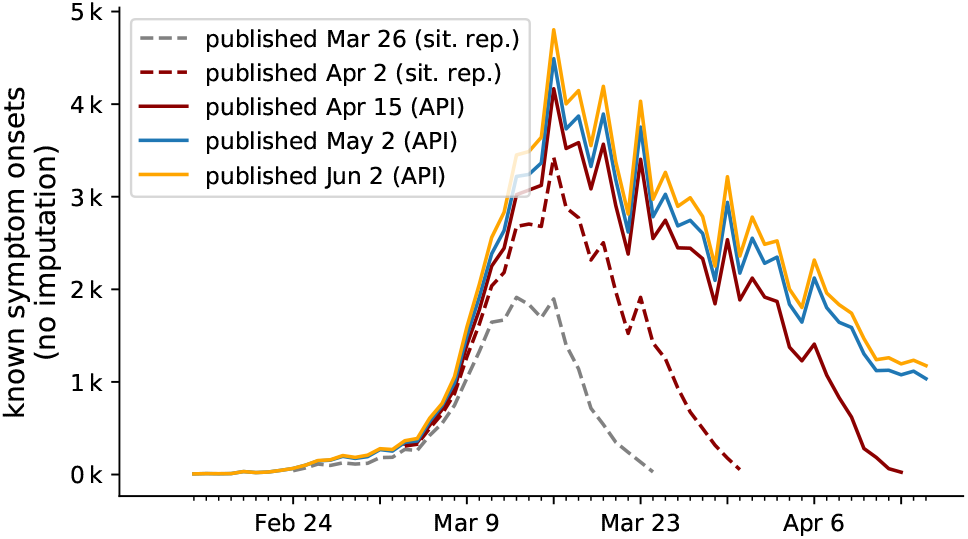
The curve of known symptom onset continuously changes over time, as the date of onset of further reported cases is obtained. Because testing confirms the onset of symptoms in the past with varying delay, the curve not only grows at its tail but over a wide time period with each new publication. **Known** onsets are reproduced from the RKI’s daily situation reports (Mar 26 and Apr 2, read of from respective plots) and the publicly available RKI-database (Apr 15, May 2 and Jun 2). Unknown onsets of symptoms, which account for 40% of total number of cases, are not considered here. Hence the curve on displey here is not the full epi curve.

During the initial outbreak of a disease, it is common that only very preliminary data and statistics on testing is available. This was also the case at the time of writing of our initial manuscript [1]. Since then, several improvements of the available data were implemented. Improvements include details such as testing statistics, but also an estimate of the epi curve (the number of cases based on the date of symptom onset) via imputation and Nowcasting. For the epi curve, complete data on symptom onset is only available for 60% of cases, and the remaining 40% of onsets need to be imputed based on the reporting date[19]. Fortunately, the publicly available RKI database contains both date of onset of symptoms and reporting for individual cases and thus implicitly also the date of testing, which in general is one day earlier than the report. Now, with new data, we come to the conclusion that reported case numbers — although they might derive from variable testing — are still useful to infer the actual disease dynamics. As we will demonstrate below, our major conclusions remain unchanged.

In particular, evidence for the key characteristics of the first wave — strong exponential growth in new cases, change in transmission dynamics over a limited time period and slow exponential decline — can be derived from the available data, even if changes in testing are considered.

**To investigate the impact of testing, we first focus on two central quantities:** i) the number of tests that are performed, say, on a given day or in a given week and ii) the fraction of the performed tests that are positive — a positive tests translates to a confirmed case.

Let us consider two simple limiting cases, in which only one of these quantities changes and the other one remains constant: If a constant number of tests is performed day-after-day and we observe a growing fraction of positive test results, this corresponds to an increase of the underlying case numbers. Conversely, if the number of tests is increased and we find a constant fraction of positive tests, this also implies an increase of the underlying case numbers^2^. Fig. 11 A, B shows that in Germany in early March both, the number of tests as well as the fraction of positives increased simultaneously. This simultaneous increase indicates a significant growth in new case numbers.

**FIG. 11.**
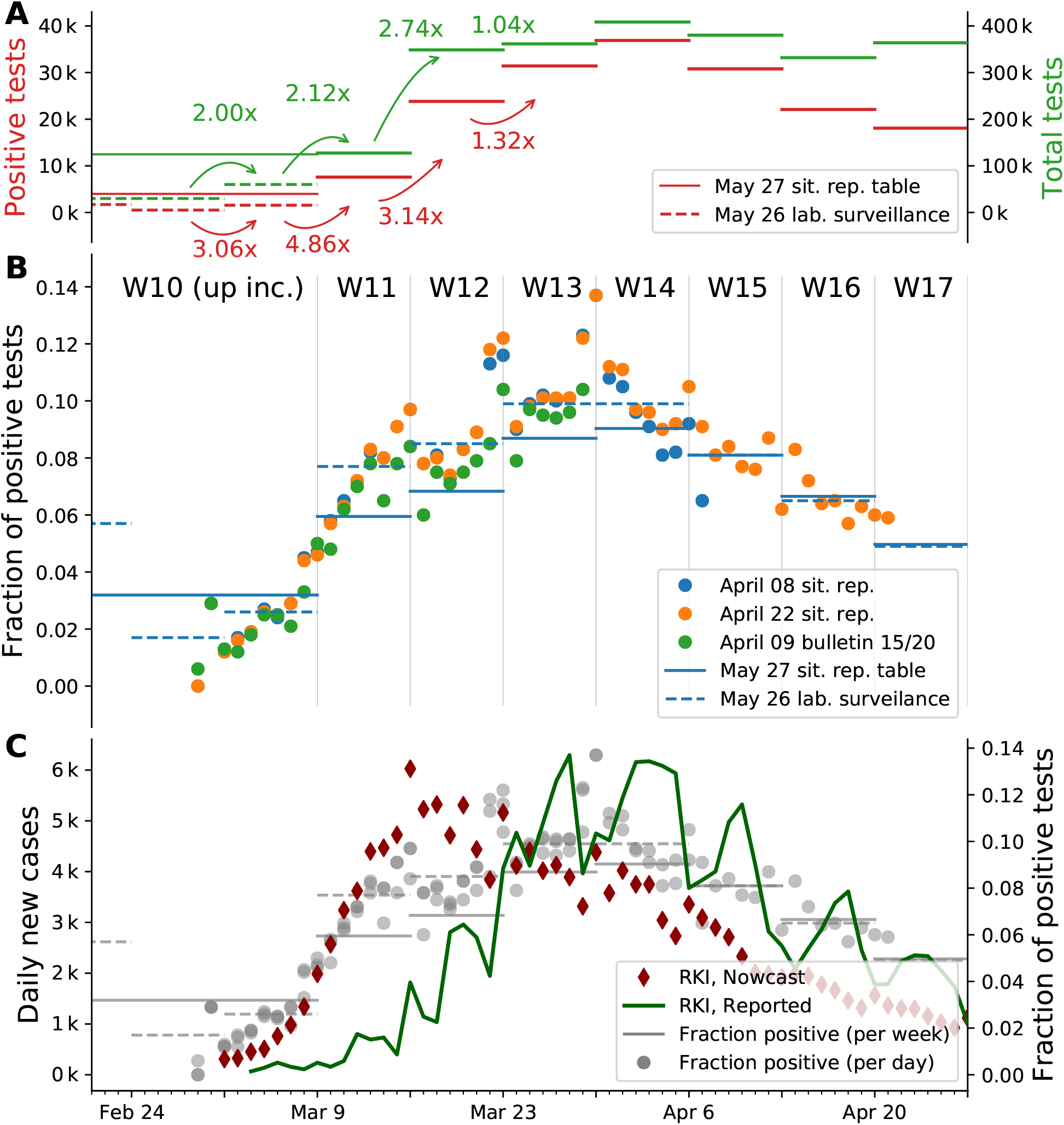
Weeks 10 to 12 show strong growth in in the number of new cases, which was not limited by the early testing capacity. **A:** Comparison of number of positive test results with the number of tests performed for each week. Reproduced from Table 5 in [21] and extrapolated from [22]. Note: Numbers for week 10 and earlier are represented by a single data point in the first source and individually in the latter. The week-over-week increase uses available weekly data. **B:** Mid-term changes in the fraction of positive tests is more obvious in the daily data (points) than in the weekly (bars), especially in early March. Daily values are taken from situation reports [21, 23, 24] (full dataset) and the epi bulletin [22, 25] (ARS dataset). Weekly values, represented as horizontal lines, are taken from a situation report table and a weekly lab surveillance report (ARS dataset). Note: the latter represents a subset of all tests. Compared to the situation report, the ARS dataset lists weeks 8 to 10 individually. **C:** Overlay of Panel B with the number of cases reported per day by the RKI and the estimated epi curve (imputation and Nowcasting, as described in [19]). The fraction of positive tests correlates with the number of reported cases from week 13 onward, as the total number of tests reaches a constant level.

### A. Strong growth of new cases until week 12

By focusing on testing before week 12 in Fig. 11 A and B, we can deduce a strong growth in daily new cases, as both the fraction of positives as well as the number of performed tests rise (matching the combined two scenarios above).

For the time before week 12, the number of tests changed week-to-week and a direct link between the test fraction and the reported cases does not hold. However, we can assume a constant level of testing within one week (Fig. 8 in [21]). At the same time, we see a continuous increase in the fraction of positives within the week (Fig. 11 B). Especially going from week 11 to week 12, where we have both, an increase in testing (from week-to-week) and an overall increase in the fraction of positives (from day-to-day), this implies a strong growth of new infections.

For weeks 12 onward, the number of performed tests stays roughly constant. Thus, the fraction of positive tests directly links to the number of reported cases, and both indicate a decline in the underlying (true) case numbers that starts in week 14. This conclusion is further supported by the high level of testing that starts in week 12: Testing at a constant *and* high level makes the fraction of positives a reliable indicator of case numbers.

**Hypothetical Scenario:** If we were to reject the above simple explanation that growing case numbers reflect growing numbers of infections, there is one alternative scenario to explain the observed trend. As this scenario has frequently occurred in the public debate on the spread of COVID-19 in Germany, we discuss it briefly.

The underlying assumption in this scenario is that the few tests that were performed during the initial outbreak until week 11 missed most of the actual cases, i.e. a large pool of infected persons would have existed unobserved. Then, at the same time at which the amount of tests was increased from week 11 to 12, coincidentally the effectiveness of the testing could have increased, so that the unobserved pool (of constant size!) is identified and, thus, apparent case numbers rise. Given the rigorous criteria (based on symptoms and risk of exposition) that were required from patients in order to qualify for one of the early tests, we deem this scenario of an unobserved and constant pool to be quite unlikely.

### B. The reporting delay relates reported cases to disease dynamics

We here focus on the disease dynamics that shape the peak of the epi curve, corresponding to the maximum new daily infections (see again Fig. 11, C, red). We notice that the increase of the fraction of positives tests (gray) continues longer and more smoothly than the increase in the epi curve (red). Thus, in the following, we discuss that testing from week 12 on reliably describes the epi curve in weeks 11–13. In general, we find as a rule of thumb that the majority of positive tests of week *i* have onsets in week *i −* 1.

The key is the connection between the date of symptoms onset (when symptoms first show), the testing (when the symptom onset is confirmed or an asymptomatic case is uncovered), and the reporting date (when a positive test-result is registered). Any reported case must inherently be preceded by a test and according to the RKI, positive test results are reported within 24 hours to the responsible health department. Thus, the date of testing is taken as the day before reporting in the rest of the analysis. The remaining task is to reveal the connection between symptom onset and reporting date, i.e. the reporting delay for each individual case.

In Fig. 12, we detail the reporting delay by plotting distributions of *how many days after the symptom onset a case is reported*. For example, if each and every infected person would receive a test result (become a reported case) exactly three days after they showed symptoms, then the plotted distributions would have only one entry: a delta-peak at three days. However, we see that most reports arrive 1–7 days after symptom onset, where the details of the (lognormal) distribution depend on the week of onset of symptoms.

**FIG. 12.**
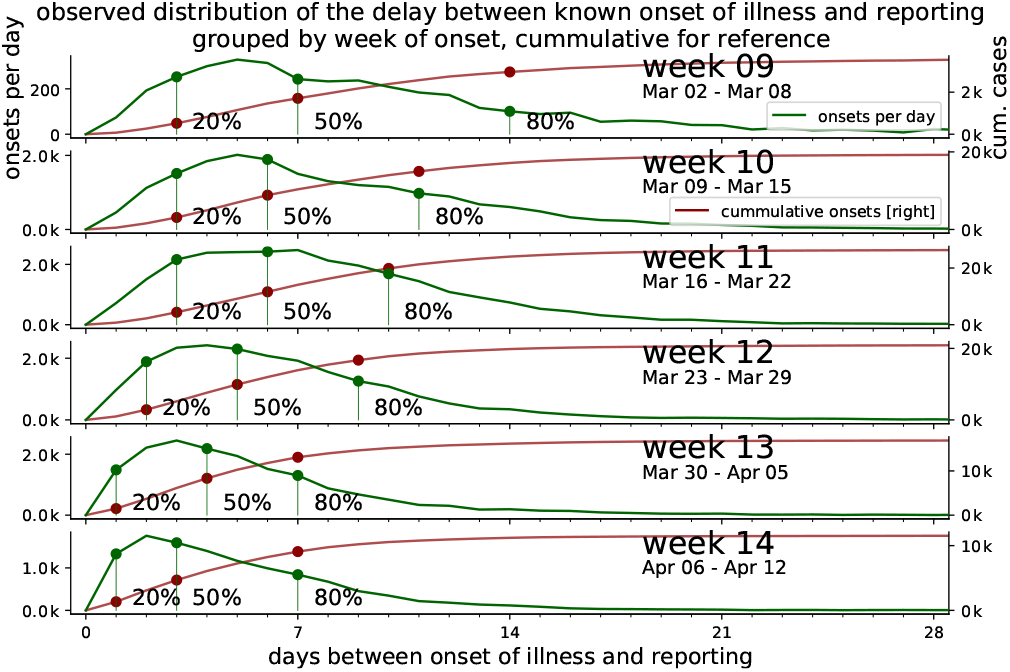
The onsets of symptoms are confirmed by testing at a later and varying point in time, which accounts for most of the delay until all or the main fraction of known onset of symptoms *(IstErkrankungsbeginn* in RKI-database) are reported. From the RKI data, the number of cases per delay between onset of illness and reporting (i.e. *RefDatum* and *Meldedatum)* for cases with known onset of symptoms *(IstErkrankungsbeginn)* are counted for each week. The fraction of reported cases out of the total onsets up to a delay are highlighted for 20%, 50% and 80%. The cumulative number of cases reported up to each delay is displayed for reference.

Heavy tails in the distributions correspond to long reporting delays. Until and including week 12, the distributions have heavy tails. After week 12, the distributions have lighter tails. This provides some intuition of the distributions and the meaning of the heavy tails: *most* of the symptom onsets are reported within the first week but *some* will be reported much later, so that the shape of the distribution still keeps changing. If the test level is low, *more* cases will be reported later and the tails of the distribution are heavier. This latter effect is what we see for the onsets during the first weeks until 11; due to limited testing capacities, many cases were only reported weeks later — once more testing was available.

The distributions of the reporting delay give information about how timely the reporting is, on average (Fig. 13). Focusing on week 11, 20% of all the onsets of symptoms that were found to be in this week were reported very quickly, within 2.5 days (blue dashed line). Within 5 days, half of all onsets have been reported (red solid line) and within 9 days, the fraction of onsets from this week that have been reported rises to 80% (blue solid line). As a practical example, let us look at the onsets that occurred on Wednesday of week 11: Half of all onsets get reported very quickly, until Sunday, and the remaining half is only reported over the following weeks.

**FIG. 13.**
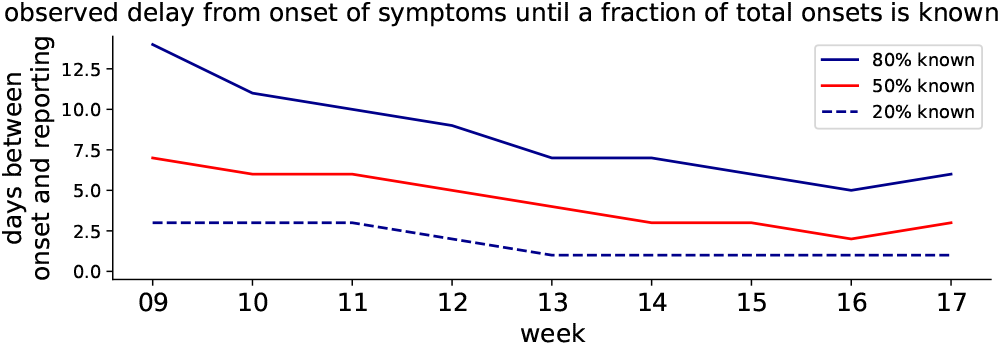
The reporting delay decreases from week 9 to week 14. Grouped by week, the delay between onset of symptoms and the reporting of 20%, 50%, 80% fractions of all known onsets is shown (cf. dots in Fig. 12).

This example also hints at a dependence of the reporting delay on the weekday. Clearly, less tests are performed during weekends. Hence, if a symptom onset occurs on Monday, it is more likely to be tested and reported within the same week than if it occurs on Friday. For later days of the week, the fraction of tests (and cases) that is performed (and reported) not in the same week but only in the next week rises systematically.

The shape of the distributions (Fig. 12) and the weekday-dependence motivate the rule of thumb mentioned earlier: 80% of all the symptom onsets that occur in a given week are reported by the end of the following week. However, due to the weekday-dependence, only around half of all onsets are found within the same week — much less during weeks 9–11, when testing was at capacity limits. In conclusion, high test levels in week *i* give confidence in the epi curve of week *i −* 1.

### C. Decomposing the epi curve into weeks of testing

Having established the delay between symptom onset and reporting, we can decompose the epi curve and identify parts of the curve that stem from certain weeks of testing. We do so by reconsidering the reporting delay. We may ask: *Given the test results of a chosen week, how are the dates of symptom onset that we found in the chosen week distributed over the previous weeks?*

In Fig. 14 A, B we collect all the symptom onsets that were found by testing in week 12 (blue), in week 13 (orange) and in both weeks combined (dashed). As we see, the peak of the full epi curve (red) on March 16 is dominantly composed of cases that were tested in week 12 and 13, weeks that already featured the high level of testing. This decomposition — which part of the curve stems from which tests — further confirms what we saw earlier: high testing in a week gives confidence in the epi curve of the previous week.

**FIG. 14.**
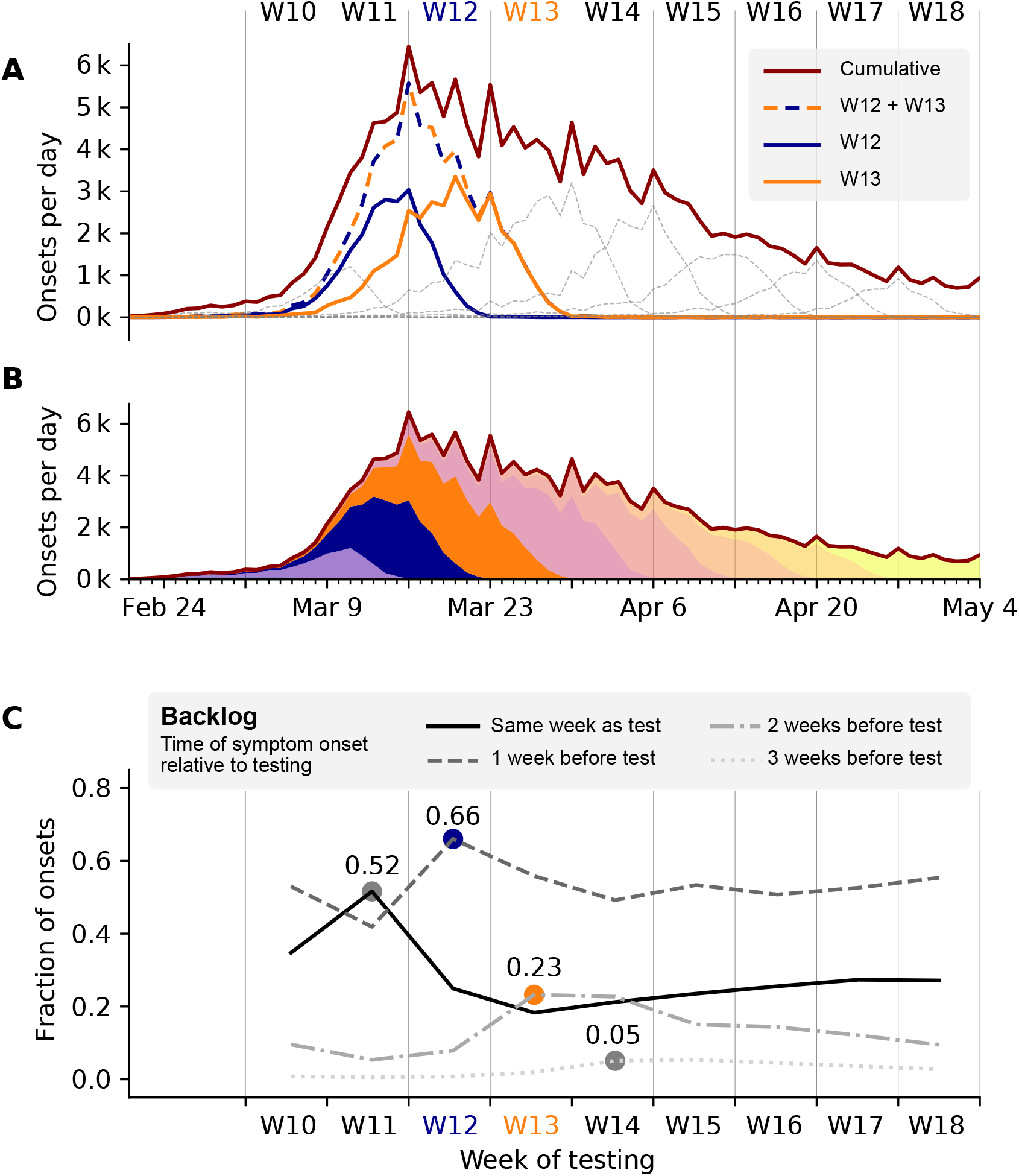
Testing in one week confirms onsets of symptoms that occur up to 4 weeks earlier. The extend of this effect is analyzed based on the RKI database through decomposition by allocation of onsets of symptoms to weeks of testing. It is assumed that the delay between the time of testing and *Meldedatum* is 1 day. Tue-Mon *Meldedatum* is taken as a proxy for Mon-Sun testing. **A** Onsets of symptoms per day curves allocated to weeks of testing, weeks 12 and 13 are highlighted. Most known onsets around the peak of the epi curve in week 11 are confirmed by the testing in weeks 12 and 13. **B** stacked decomposition of the epi curve into weeks of testing. **C** To reveal crucial information about week-to-week change in the number of total onsets based on one week of testing, the shape of the distributions of onsets of symptoms confirmed by that week of testing is characterized. The fraction of onsets in the same week and each preceding week out of the total onsets confirmed by the week of testing is calculated. This indicates, the portion of a week’s positive tests confirming onsets in the same week or in preceding weeks (max. 3 weeks earlier). The evolution of these 4 values is plotted by the week of testing. The peak of the epi curve can be tracked through testing results of weeks 11 to 14 as a maximum in the same-week/n-weeks earlier fraction of onsets confirmed in those respective weeks: 52% of all cases confirmed through testing in week 11 had onset of symptoms in the same week. Even more notable: 66% of positive tests in week 12 are linked to onsets 1 week earlier: in week 11. For comparison, see Fig. ??

With the decomposition of the epi curve at hand, we may pick one particular week of testing and compare the number of onsets in different weeks that were confirmed in the testing-week we picked. In other words, we are interested in the *distribution of onsets per week seen by the testing in one single week*.

As viewed from one single week of testing, we distinguish four categories according to the delay between onset and testing (Fig. 14 C): onsets in the same week as the test (solid), onsets one week earlier than the test (dashed), onsets two weeks earlier (dash-dotted), and onsets three weeks earlier (dotted). By comparing the fraction of cases in these categories week-over-week, we can reveal the *backlog of testing*. The backlog of testing corresponds to the last three categories; it describes how many cases were not tested within the same week (different dashing). Looking at the backlog week-over-week helps us to identify weeks during which the limit of testing capacity might have been reached or the testing policy might have changed.

When considering the respective maxima of the backlog-categories (colored dots in Fig. 14), we find that backlog was build up especially during week 11. In week 11, most onsets stem from the same week (52%, maximum of the solid line). At the same time, in week 11 there was very little backlog; only few cases from previous weeks were found (minima of the dashed lines). In week 12, we find that most cases stem from the previous week — namely week 11 (66%, maximum of the dashed line). This trend continues in weeks 13 and 14, which exhibit comparably high fractions of onset 2 weeks and 3 weeks earlier, respectively, each pointing to week 11 (maxima marked by dots). Together, this (self-consistently) supports the strong growth of new onsets especially during week 11; a strong rise of cases before week 11 is less likely because it did not manifested in the backlog.

### D. Available data on testing

The epi bulletin [26] outlines the different networks that the RKI uses to source information on testing: *Voxco, RespVir*, the antibiotics-resistance-surveillance (ARS) [22] and lab-accociation queries. These sources are compiled into weekly data-sets with total number of tests and positive tests, which are published in the daily situation report once a week.

Data from the ARS contains daily number on testing and a separate weekly report is published on the RKI website. The ARS dataset covers 25–30% of the total number of tests reported by the RKI, as only 62 of 180+ labs participate. The ARS data-set shows a mean delay between sampling and testing between 1 and 1.2 days except for weeks 12 to 15, where the delay is 1.5 days, peaking in week 13 at 1.8 days.

An overview of all publicly available data on testing for march 2020 is presented in Fig 11. The following observations along with additional comments are based on this presentation:

- From week 8 to week 12 the number of tests rises week to week by a factor greater than 2. 120k is a combined number for weeks up to 10. Individual numbers of tests for those weeks has to be estimated with help from the ARS-subset (Fig. 11 B *May 26 lab. surveillance)*. Assuming ARS is representative the number of test performed in week 10 should be around 60k, 30k in week 9 and 30k in all weeks up to and including 8, extending the exponential pattern.
- The number of tests remains on a high level from week 12 on. In the range of 340–430k.
- The number of positive tests rises faster than the total number of tests until week 14.
- The fraction of positive tests per week peaks around 10%, relatively low compared with neighbouring countries.
- The fraction of positive tests per day varies with time from 2% around March 1 to around 10% in weeks 13 and 14, peaking at 14% at the end of March. Afterwards declining to less than 2% in week 20 (not shown in figure). The day-to-day rise in week 10 and 11 is more pronounced than the weekly average would suggest.
- The increase in the fraction of positive tests does not correlate to the rise in number of reported cases until week 13, but correlates with the decline in reported cases from week 13 on, which is expected as the total number of tests fluctuates around 380k tests per week on a high level.
- The ARS data shows a steady day to day increase in positive fraction of test in weeks 10 and 11. Weekends show a higher fraction, while the total number of tests is lower (daily total number not shown in the figure). Deviating from the rise in the positive fraction, weeks up to 8 have a 3 times higher fraction of positive results than week 9.
- The maximum test-capacity per week as reported by the labs increased to 1M in week 19, showing strong growth until week 14. A week to week doubling in test capacity continues for two more weeks compared to growth in number of tests performed (not shown).

For the total data-set, the fraction of positive tests varies from 1.5 to 7.2% for different states. Not a single day of testing for individual states exceeded 20% positive results.

## VI. SUMMARY & CONCLUSIONS

In these technical notes, we have comprehensively addressed questions and comments regarding our recent publication [1]. First, we compared direct, model-free estimates of the reproduction number to the ones obtained from dynamical modeling. To this end, we established synthetic ground-truth data based on an SIR model and subsequently inferred the reproduction number based on various complementary approaches that are in practical use. We revealed how sudden changes in the spreading rate — as expected from the broad and swift implementation of non-pharmaceutical interventions and concurrent changes in behavior — can lead to counter-intuitive transient drops in new reported cases. Most importantly, we found that modeling of spreading dynamics can correctly capture effects of sudden changes in the spreading rate.

Second, we provided extensive background on our modeling rationale, which combines differential-equation based modeling of dynamics with Bayesian parameter inference and formal model comparison. Within the Bayesian framework, we argued that based on prior knowledge, the most plausible models explaining the data can be systematically identified and also updated as new information becomes available. We also discussed why we do not think that strong effects of herd immunity are plausible given our present knowledge.

Third, we analyzed additional data on the SARS-CoV-2 spread in Germany, which has become available since the completion of the analysis presented in [1]. Most importantly, we included data sets from the German Robert Koch Institute based on the reporting date as well as based on the onset of symptoms (epi curve). We analyzed the data in the framework of SIR and SEIR models, and we also tested a broad range of varying prior assumptions. We found our central results to be robust across these varying modeling assumptions and data sets, and to support the conclusions drawn in [1]. In turn, this lead us to conclude that under the conditions comparable to those in Germany, models based on reporting date are a viable alternative for analyzing the early stages of a disease outbreak, before the epi curve becomes available — as long as the reporting delay is properly modeled.

Finally, we addressed the issue of changes in the testing capacities and procedures over the course of our analysis. Most importantly, we found that, while data from the initial onset of the pandemic is presumably affected by a rise in test capacities, the crucial part of our analysis is based on a regime of comparably stable testing. In particular, we concluded that the inference of the second and third change point is widely unaffected by testing.

Overall, the analysis here evaluated the robustness of our previously reported results with respect to statistical and dynamical modeling assumptions as well as complementary data sources and provided additional support for the central conclusions of our publication [1]:

1. combining epidemiological modelling with Bayesian inference enables a robust assessment of the spreading of infectious diseases in a timely manner;
2. the spreading dynamics can only be inferred with a considerable delay (due to incubation periods and testing/reporting delays);
3. applied to the COVID-19 outbreak in Germany, it appears most plausible that all interventions together with the concurrent change in behavior reduced the effective growth rate *λ^∗^*, and that *λ^∗^* dropped substantially below zero close to the time of the third intervention.

## Data Availability

We provide the code for generating graphics and all the different analyses included this manuscript and its supplementary materials at https://github.com/Priesemann-Group/covid19_research/tree/master/Technical%20notes%20on%20Dehning%20et%20al.%2C%20Science%2C%202020

## SUPPLEMENTARY INFORMATION: FIGURES

**FIG. 15.**
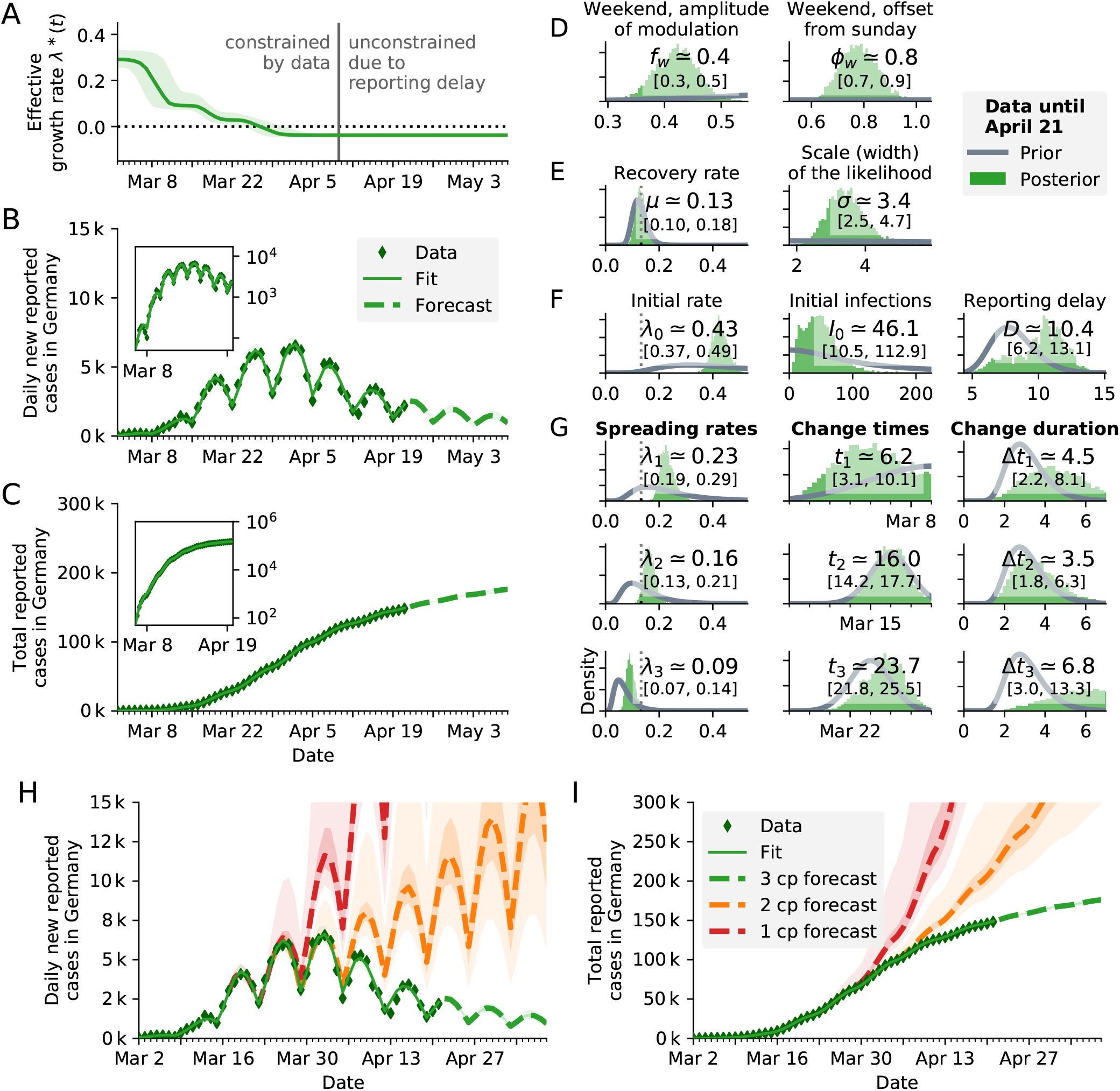
SIR model. (see Fig. 3 of [1]) using the reporting date (Meldedatum) of the RKI data for inference. **A:** Time-dependent model estimate of the effective spreading rate *λ^∗^(t)*. **B:** Comparison of daily new reported cases and the model (green solid line for median fit with 95% credible intervals, dashed line for median forecast with 95% CI); inset same data in log-lin scale. **C:** Comparison of total reported cases and the model (same representation as in B). **D–G:** Priors (gray lines) and posteriors (green histograms) of all model parameters; inset values indicate the median and 95% credible intervals of the posteriors. **H–I:** The fitted model with two alternative forecasts. We consider in addition one scenario where only one intervention happened (red) and one where two interventions happened (orange). Includes 50% and 95% CI.

**FIG. 16.**
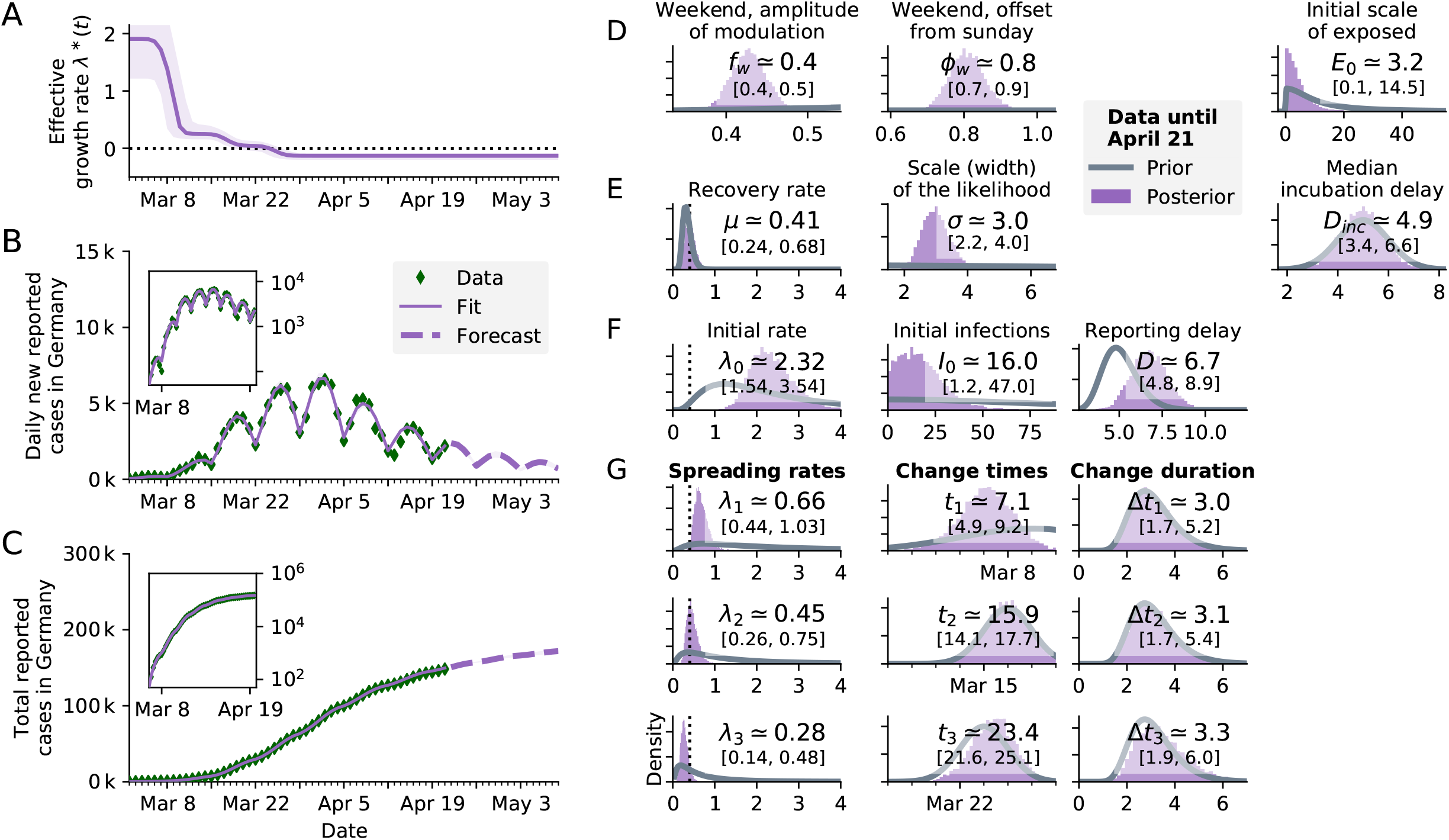
SEIR-like model. (see Fig. S3 in Supplementary Information of [1]) using the reporting date (Meldedatum) of the RKI data for inference. **A:** Time-dependent model estimate of the effective spreading rate *λ^∗^(t)*. **Note:** Due to different model dynamics, *λ^∗^(t)* can only be compared *qualitatively* between SEIR and SIR models. The numeric values of the rates *(µ, λ* etc.) differ between models because they reflect the duration a person remains in a given compartment. **B:** Comparison of daily new reported cases and the model (purple solid line for median fit with 95% credible intervals, dashed line for median forecast with 95% CI); **inset** same data in log-lin scale. **Note:** We currently do not (yet) incorporate the uncertainties that are introduced by nowcasting, compared to using the reported cases. This leads to *over-confident* parameter estimates, including the effective spreading rate *λ^∗^(t)*; the shown uncertainties are underestimated. **C:** Comparison of total reported cases and the model (same representation as in B). D–G: Priors (gray lines) and posteriors (purple histograms) of all model parameters; inset values indicate the median and 95% credible intervals of the posteriors.

**FIG. 17.**
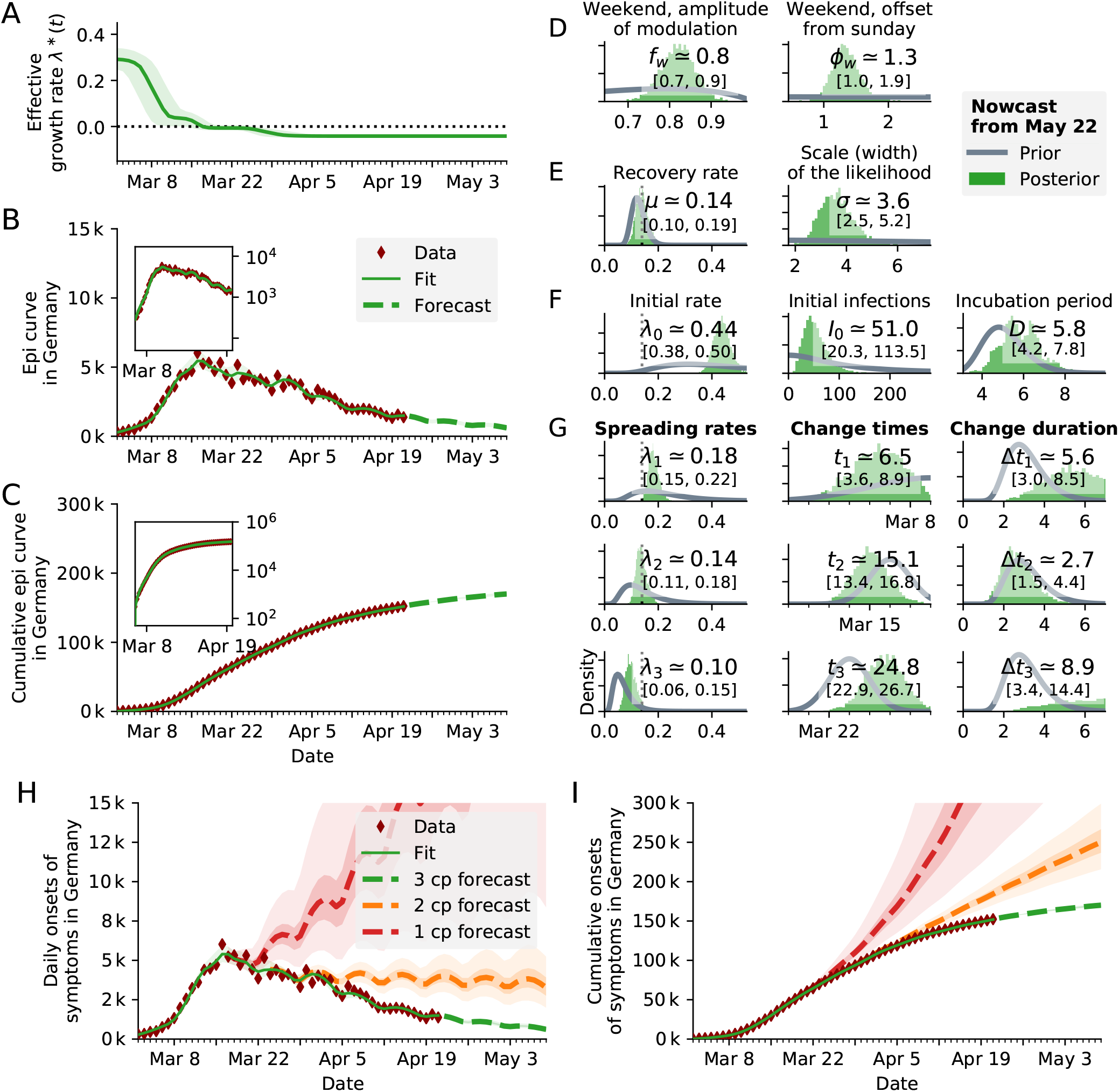
SIR model using the onset of symptoms. (unsmoothed Nowcast from May 22 [16]) of the RKI data for inference. The median of the lognormal prior of the delay between infection and onset of symptoms has been set to 5 days (right-most panel F). **A:** Time-dependent model estimate of the effective spreading rate *λ^∗^(t)*. **Note:** We currently do not (yet) incorporate the uncertainties that are introduced by nowcasting, compared to using the reported cases. This leads to *over-confident* parameter estimates, including the effective spreading rate *λ^∗^(t)*; the shown uncertainties are underestimated. **B:** Comparison of daily new reported cases and the model (green solid line for median fit with 95% credible intervals, dashed line for median forecast with 95% CI); inset: same data in log-lin scale. **Note:** We currently do not (yet) incorporate the uncertainties that are introduced by nowcasting, compared to using the reported cases. This leads to *over-confident* parameter estimates, including the effective spreading rate *λ^∗^(t)*; the shown uncertainties are underestimated. **C:** Comparison of total reported cases and the model (same representation as in B). **D–G:** Priors (gray lines) and posteriors (green histograms) of all model parameters; inset values indicate the median and 95% credible intervals of the posteriors. **H–I** The fitted model with two alternative forecasts. We consider in addition one scenario where only one intervention happened (red) and one where two interventions happened (orange). Includes 50% and 95% CI.

**FIG. 18.**
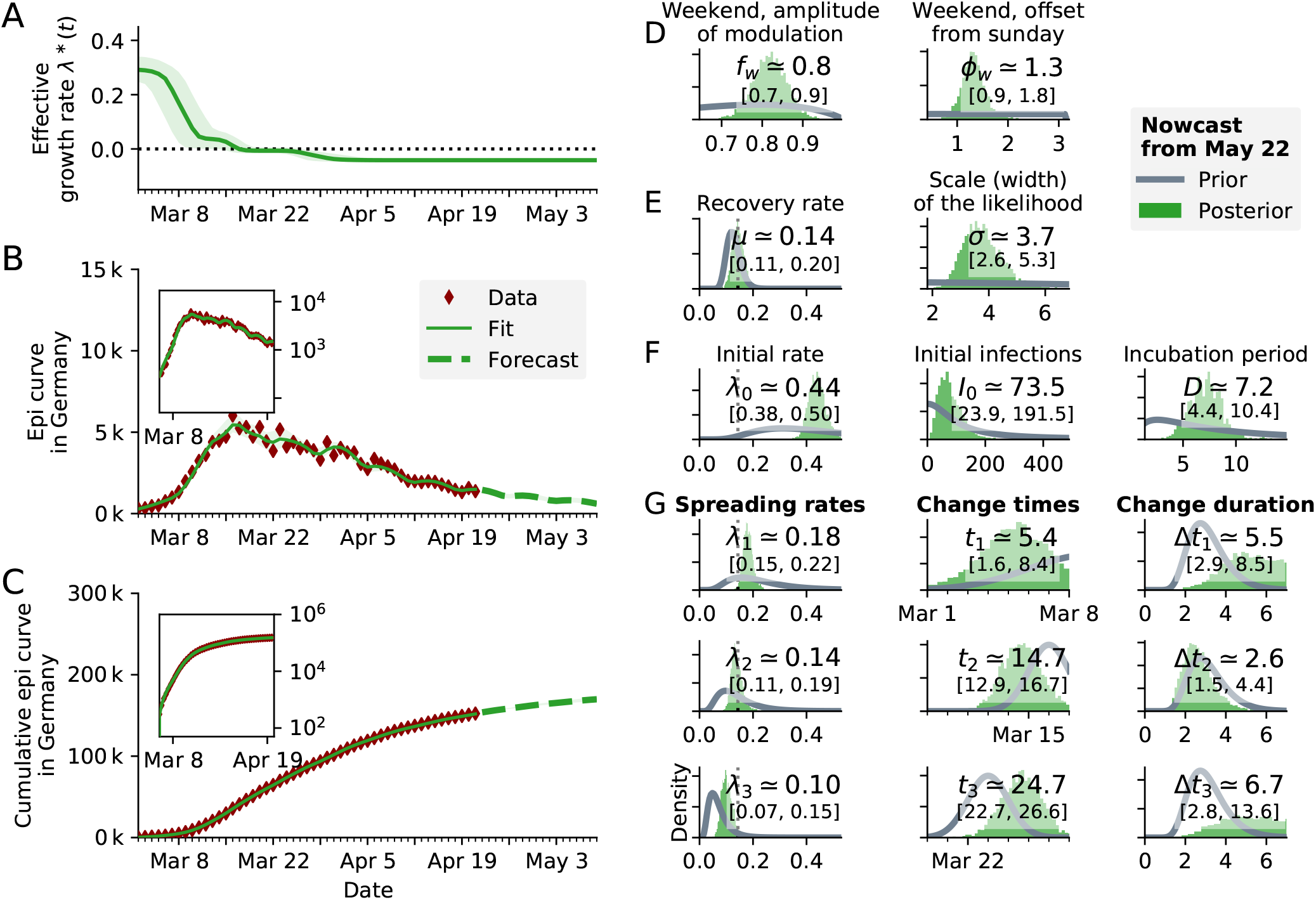
SIR model using the onset of symptoms (unsmoothed Nowcast from May 22 [16]) of the RKI data for inference. The median of the lognormal prior of the delay between infection and onset of symptoms has been set to a relatively uninformative prior. (right-most panel F). The posterior of the delay has as median 7.2 days, which is close to the expected incubation period of 5 days. **A:** Time-dependent model estimate of the effective spreading rate *λ^∗^(t)*. **Note:** We currently do not (yet) incorporate the uncertainties that are introduced by nowcasting, compared to using the reported cases. This leads to *over-confident* parameter estimates, including the effective spreading rate *λ^∗^(t)*; the shown uncertainties are underestimated. **B:** Comparison of daily new reported cases and the model (green solid line for median fit with 95% credible intervals, dashed line for median forecast with 95% CI); **inset** same data in log-lin scale. **C:** Comparison of total reported cases and the model (same representation as in B). **D–G:** Priors (gray lines) and posteriors (green histograms) of all model parameters; inset values indicate the median and 95% credible intervals of the posteriors.

**FIG. 19.**
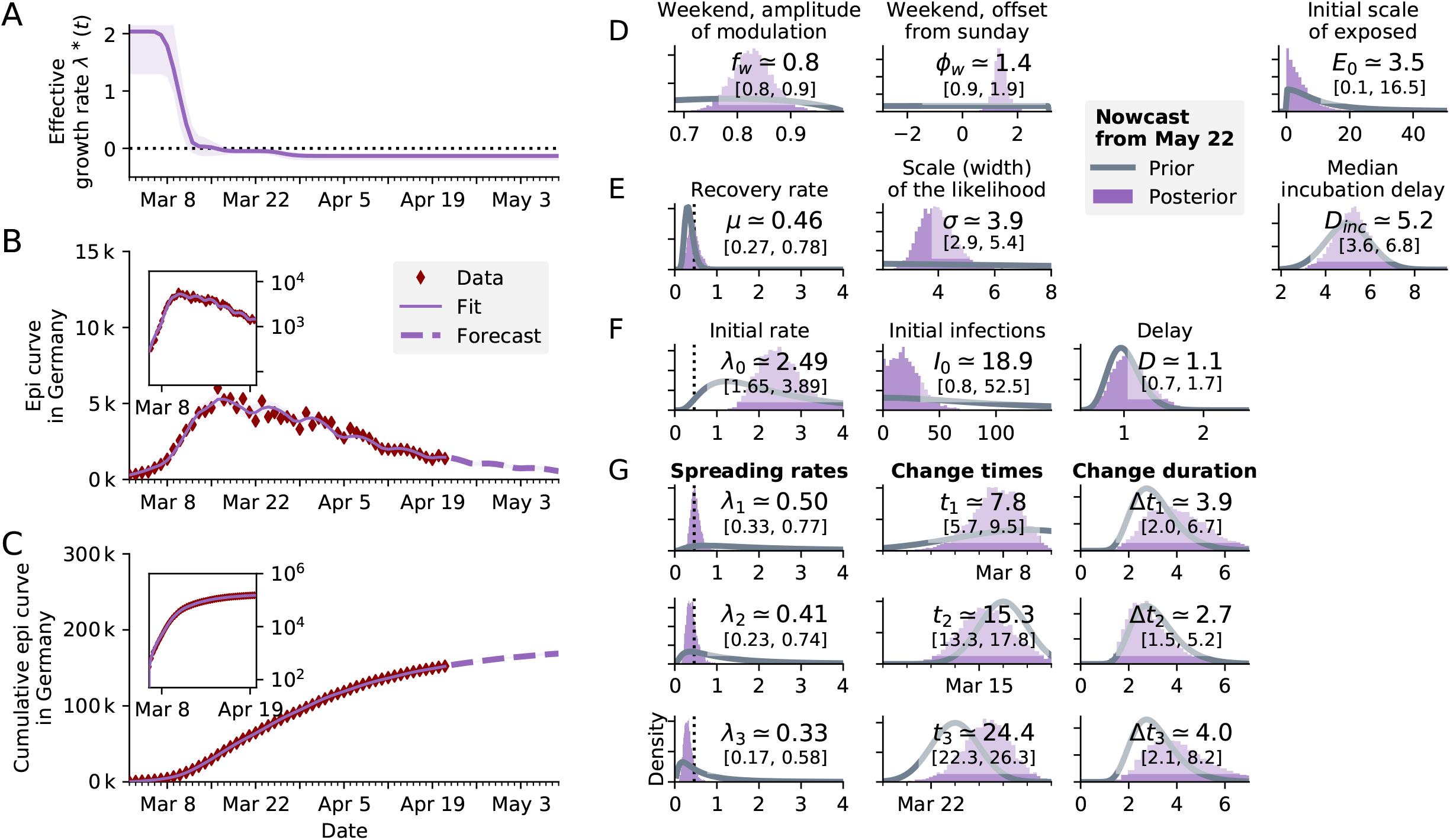
SEIR-like model using the onset of symptoms. (unsmoothed Nowcast from May 22 [16]) of the RKI data for inference. The median of the lognormal prior of the delay between infectious and onset of symptoms has been set to 1 day (right-most panel F). **A:** Time-dependent model estimate of the effective spreading rate *λ^∗^(t)*. **Note:** Due to different model dynamics, *λ^∗^(t)* can only be compared *qualitatively* between SEIR and SIR models. The numeric values of the rates *(µ, λ* etc.) differ between models because they reflect the duration a person remains in a given compartment. **Note:** We currently do not (yet) incorporate the uncertainties that are introduced by nowcasting, compared to using the reported cases. This leads to *over-confident* parameter estimates, including the effective spreading rate *λ^∗^(t)*; the shown uncertainties are underestimated. **B:** Comparison of daily new reported cases and the model (purple solid line for median fit with 95% credible intervals, dashed line for median forecast with 95% CI); inset same data in log-lin scale. **C:** Comparison of total reported cases and the model (same representation as in B). **D–G:** Priors (gray lines) and posteriors (purple histograms) of all model parameters; inset values indicate the median and 95% credible intervals of the posteriors.

**FIG. 20.**
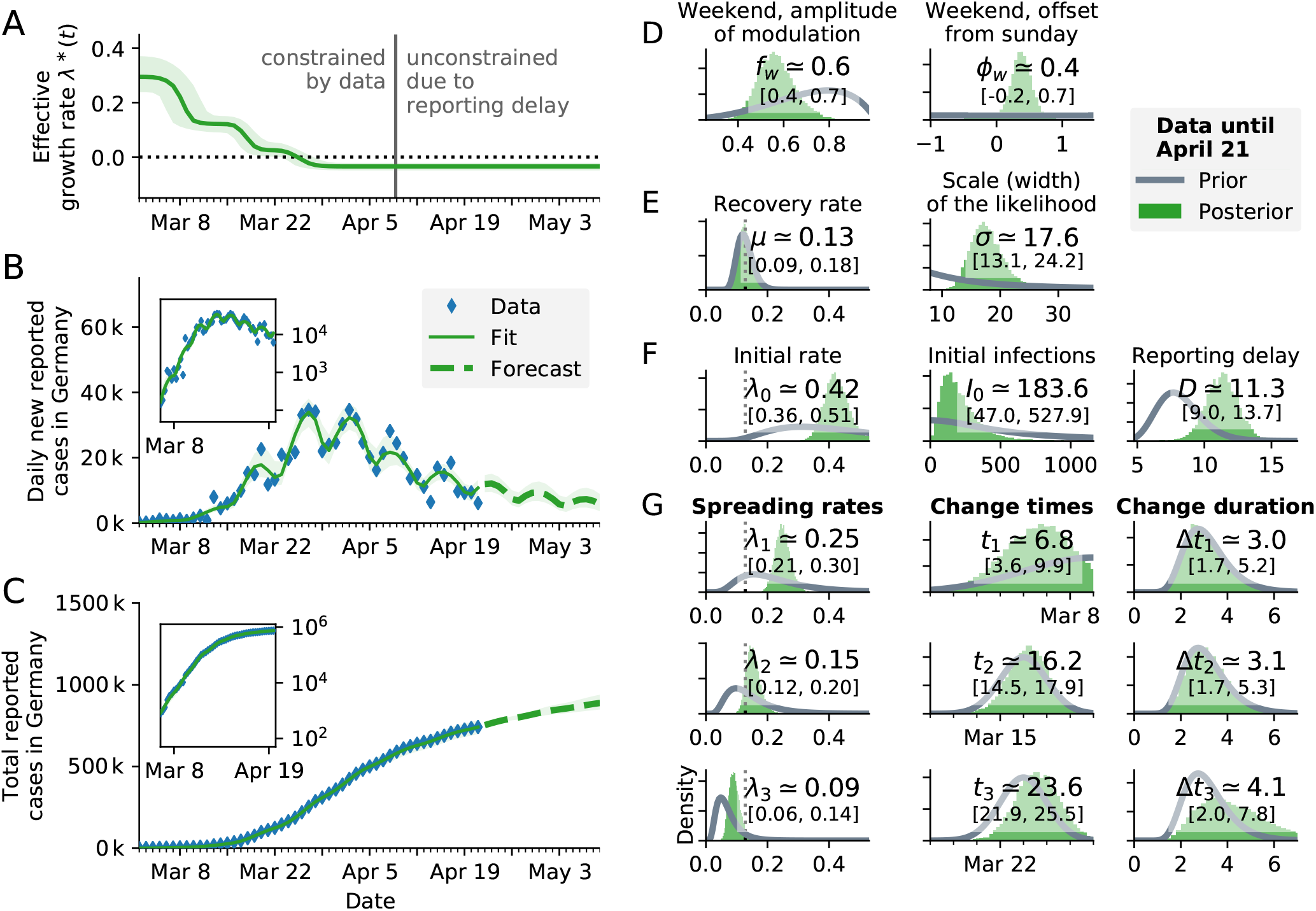
SIR model with reported case number multiplied by 5, to account for an eventual factor five of unknown cases. Results are nearly identical to original non-multiplied plot (Fig 3. in [1]), showing that a constant underreporting has a negligible effect. The median inferred spreading rates *λ* are about 0.01 larger. **A:** Time-dependent model estimate of the effective spreading rate *λ^∗^(t)*. **B:** Comparison of daily new reported cases and the model (green solid line for median fit with 95% credible intervals, dashed line for median forecast with 95% CI); inset same data in log-lin scale. **C:** Comparison of total reported cases and the model (same representation as in B). **D–G:** Priors (gray lines) and posteriors (green histograms) of all model parameters; inset values indicate the median and 95% credible intervals of the posteriors.

**FIG. 21.**
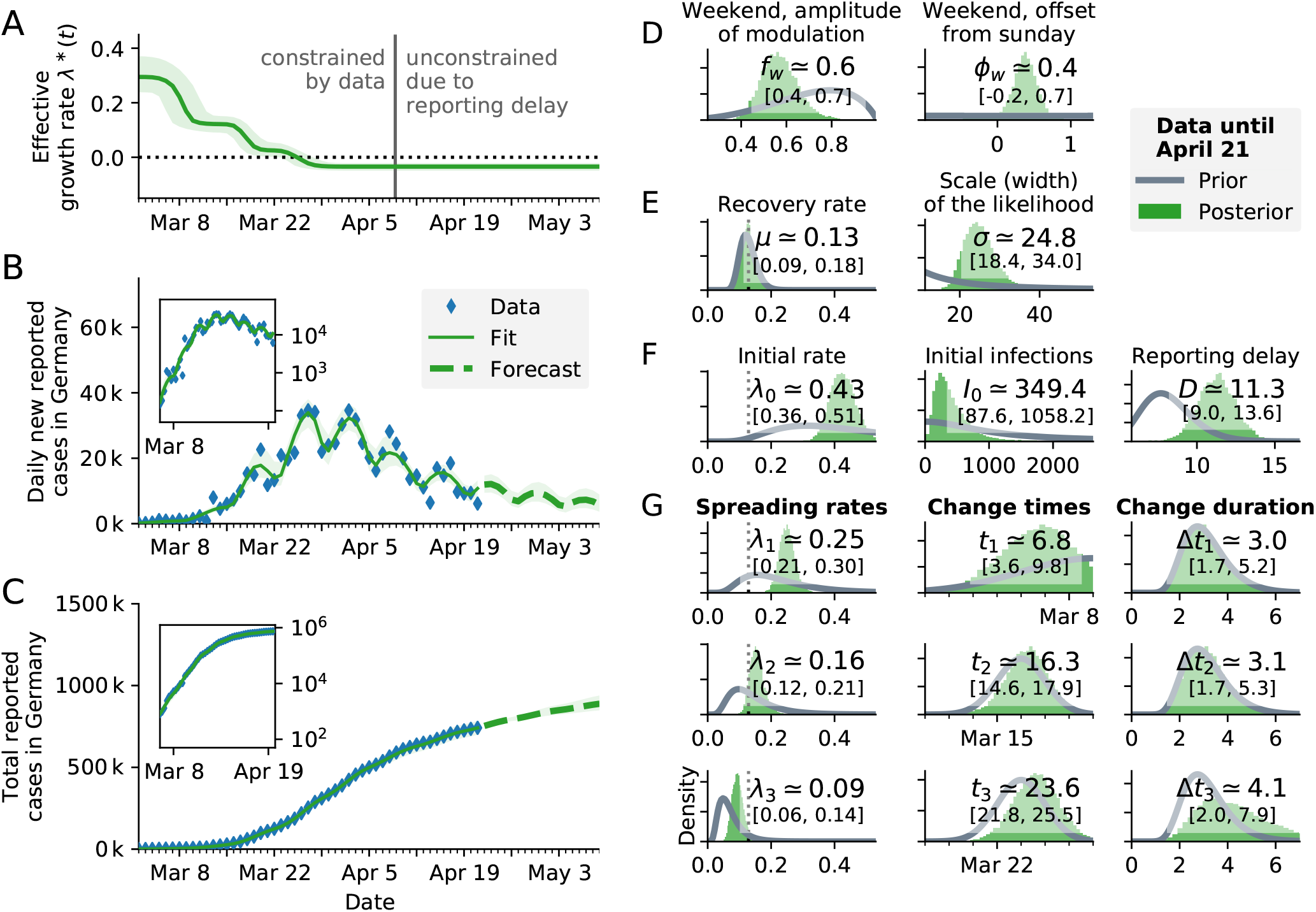
SIR model with reported case number multiplied by 10, to account for an eventual factor 10 of unknown cases. Results are nearly identical to original non-multiplied plot (Fig 3. in [1]), showing that a constant under-reporting has a negligible effect, similar to Fig. 20. The median inferred spreading rates *λ* are 0.01–0.02 larger. **A:** Time-dependent model estimate of the effective spreading rate *λ^∗^(t)*. **B:** Comparison of daily new reported cases and the model (green solid line for median fit with 95% credible intervals, dashed line for median forecast with 95% CI); inset same data in log-lin scale. **C:** Comparison of total reported cases and the model (same representation as in B). **D–G:** Priors (gray lines) and posteriors (green histograms) of all model parameters; inset values indicate the median and 95% credible intervals of the posteriors.

**FIG. 22.**
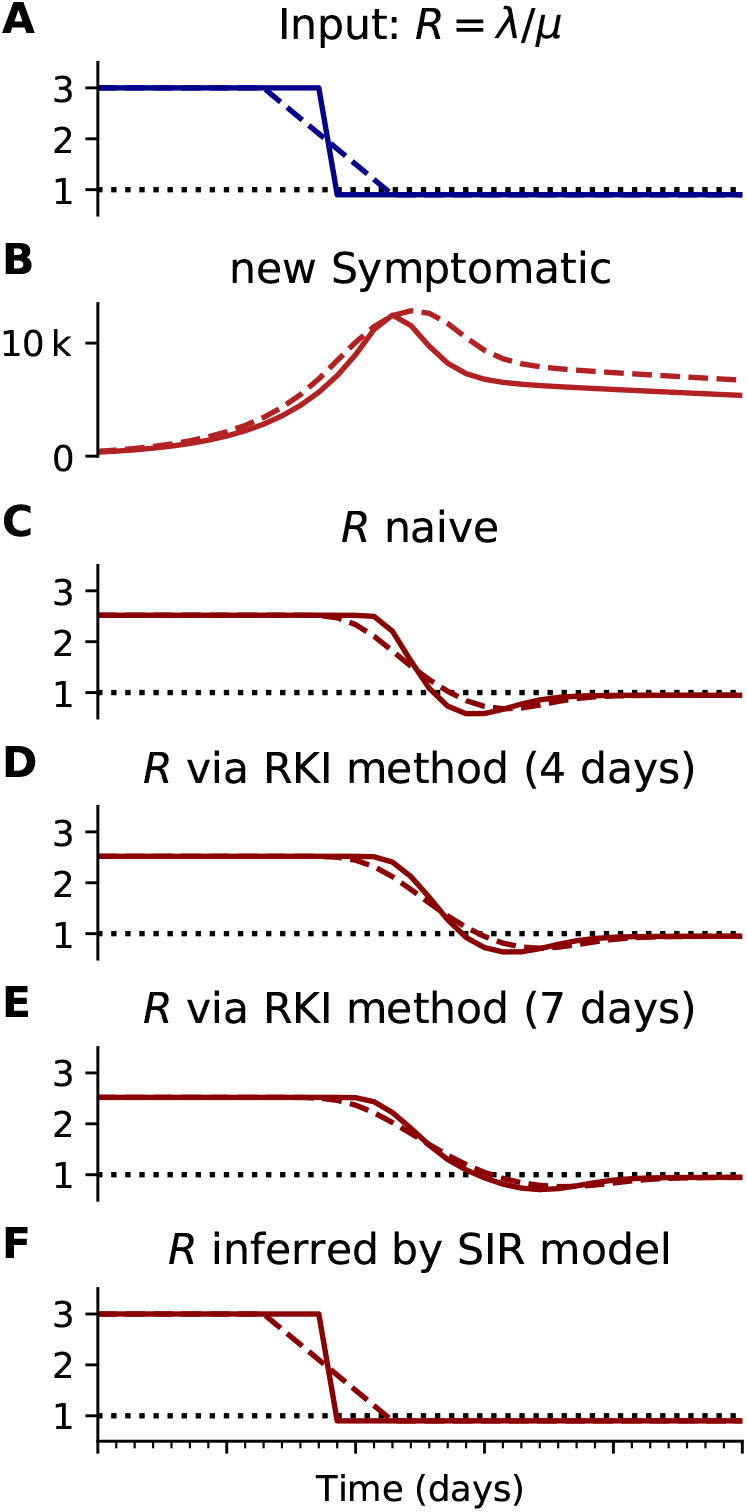
Inferring R with different methods (like Fig. 4), using a synthetic model **with** *R* = 3 to *R* = 0.9. A, **B:** Synthetic data for new symptomatic cases generated with SIR dynamics from an underlying *R* with one change point of duration 1 day (solid) or 7 days (dashed). **C:** Model-free inference of *R* based on the ratio of case numbers at time *t* and time *t − g*. **D:** Model-free inference of *R* following the Robert Koch Institute convention, i.e. using the definition of C but with averaging over a window of the past 4 days. **E:** Same as D but averaging over 7 days. Note the overlap of intervals. All the model-free methods (C–E) can show an erroneous estimate of *R* < 0.9 transiently, due to the change point in the underlying true *R*. **F:** The inferred *R* using change-point detection with an underlying dynamic model (SIR) does *not* show a transient erroneous *R* < 0.9 period. If the underlying dynamic model corresponds well enough to the true disease dynamics, then this approach reproduces the true *R* that was used to generate the data.

**FIG. 23.**
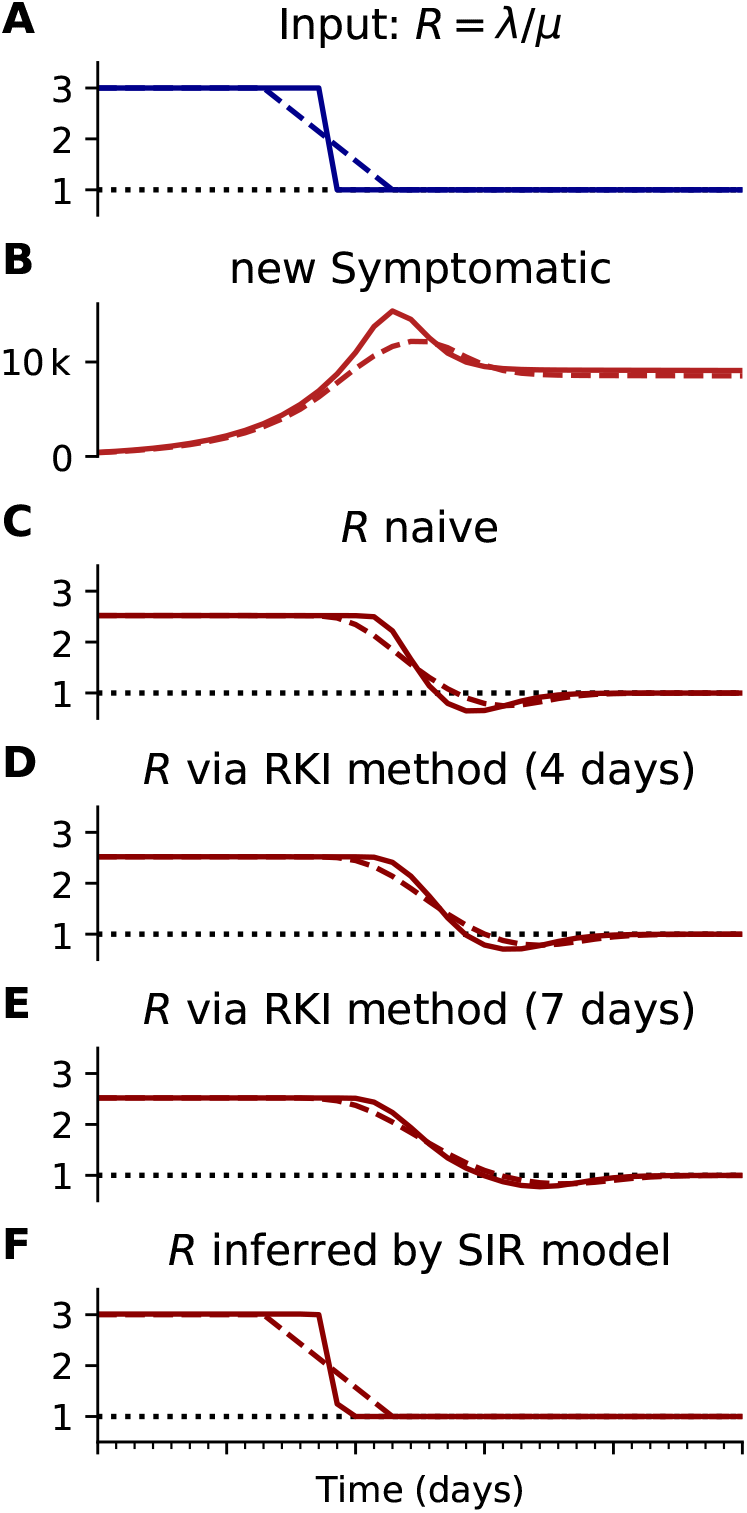
Inferring R with different methods (like Fig. 4), using a synthetic model with *R* = 3 to *R* = 1. **A, B:** Synthetic data for new symptomatic cases generated with SIR dynamics from an underlying *R* with one change point of duration 1 day (solid) or 7 days (dashed). **C:** Model-free inference of *R* based on the ratio of case numbers at time *t* and time *t − g*. **D:** Model-free inference of *R* following the Robert Koch Institute convention, i.e. using the definition of C but with averaging over a window of the past 4 days. **E:** Same as D but averaging over 7 days. Note the overlap of intervals. All the model-free methods (C–E) can show an erroneous estimate of *R* < 1 transiently, due to the change point in the underlying true *R*. **F:** The inferred *R* using change-point detection with an underlying dynamic model (SIR) does *not* show a transient erroneous *R* < 1 period. If the underlying dynamic model corresponds well enough to the true disease dynamics, then this approach reproduces the true *R* that was used to generate the data.

**FIG. 24.**
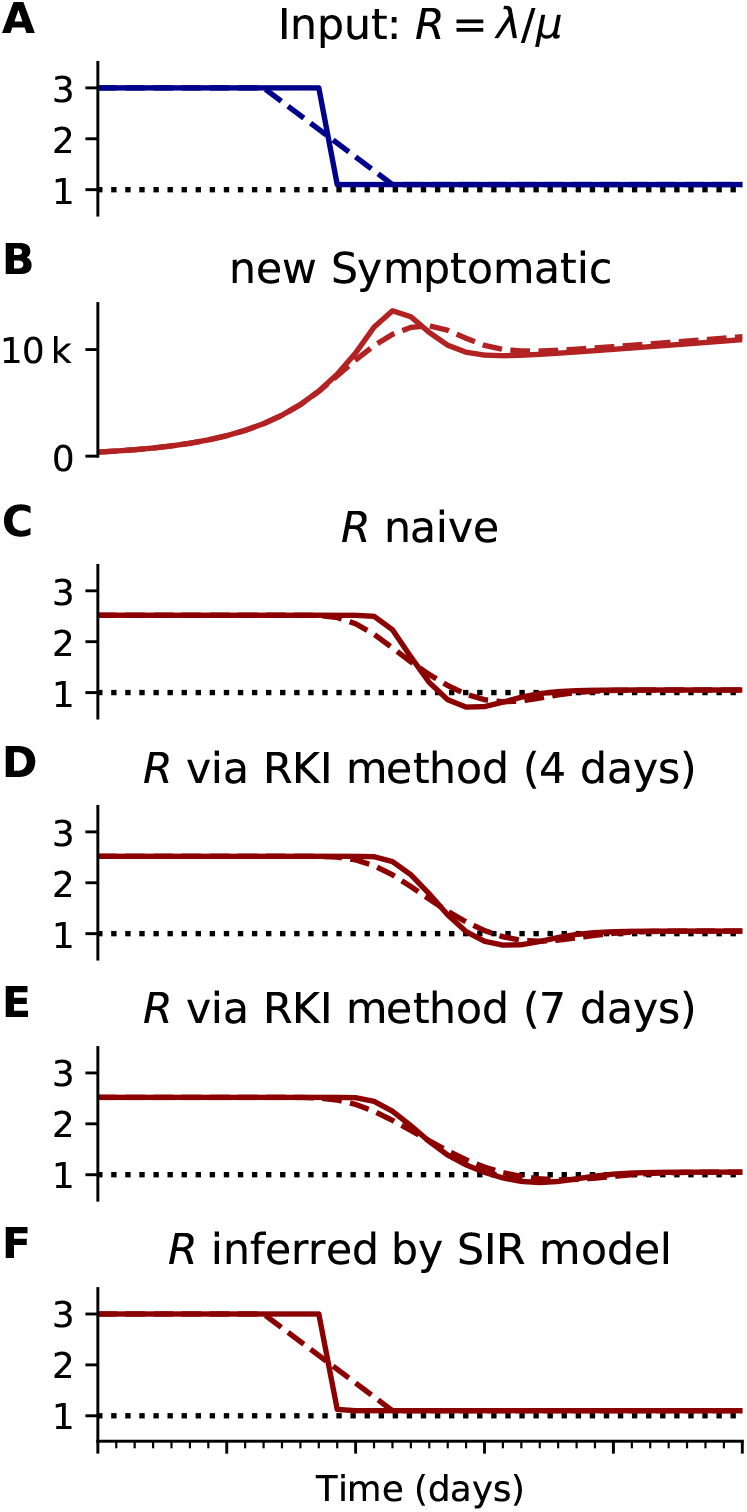
Inferring R with different methods (like Fig. 4), using a synthetic model with *R* = 3 to *R* = 1.1. **A, B:** Synthetic data for new symptomatic cases generated with SIR dynamics from an underlying *R* with one change point of duration 1 day (solid) or 7 days (dashed). **C:** Model-free inference of *R* based on the ratio of case numbers at time *t* and time *t − g*. **D:** Model-free inference of *R* following the Robert Koch Institute convention, i.e. using the definition of C but with averaging over a window of the past 4 days. **E:** Same as D but averaging over 7 days. Note the overlap of intervals. All the model-free methods (C–E) can show an erroneous estimate of *R* < 1 transiently, due to the change point in the underlying true *R*. **F:** The inferred *R* using change-point detection with an underlying dynamic model (SIR) does *not* show a transient erroneous *R* < 1 period. If the underlying dynamic model corresponds well enough to the true disease dynamics, then this approach reproduces the true *R* that was used to generate the data.

## Footnotes

1 In Germany, only for a tiny fraction of cases the reported symptom onset *(Refdatum)* is after testing *(Meldedatum)*: 1446 of 130027 cases in the RKI dataset of Jun 11.

2 The second case only holds with additional assumptions: i) the fraction of positive tests is larger than the prevalence and ii) tests are not performed randomly, both of which were met in Germany.

